# Direct Molecular Analysis of *In Vivo* and Freshly Excised Tissues in Human Surgeries with the MasSpec Pen Technology

**DOI:** 10.1101/2020.12.14.20248101

**Authors:** Jialing Zhang, Marta Sans, Rachel J. DeHoog, Kyana Y. Garza, Mary E. King, Clara L. Feider, Alena Bensussan, Michael F. Keating, John Q. Lin, Sydney Povilaitis, Nitesh Katta, Thomas E. Milner, Wendong Yu, Chandandeep Nagi, Sadhna Dhingra, Christopher Pirko, Kirtan A. Brahmbhatt, George Van Buren, Stacey Carter, William E. Fisher, Alastair Thompson, Raymon H. Grogan, James Suliburk, Livia S. Eberlin

## Abstract

Intraoperative tissue analysis is critical to guide surgical procedures and improve patient outcomes. Here, we describe the clinical translation and intraoperative use of the MasSpec Pen technology for direct molecular analysis of *in vivo* and freshly excised tissues in the operating room. In this study, the MasSpec Pen was used by surgeons and surgical staff during 100 surgeries over a 12-month period, allowing rapid detection of rich mass spectral profiles from 715 *in vivo* and *ex vivo* analyses performed on thyroid, parathyroid, lymph node, breast, pancreatic, and bile duct tissues during parathyroidectomies, thyroidectomies, breast, and pancreatic neoplasia surgeries. The MasSpec Pen enabled gentle extraction and sensitive detection of various molecular species including small metabolites and lipids using a droplet of sterile water without causing apparent tissue damage. Notably, effective molecular analysis was achieved while no limitations to sequential histologic tissue analysis were identified and no device-related complications were reported for any of the patients. Collectively, this study shows that the MasSpec Pen system can be successfully incorporated into the operating room, allowing direct detection of rich molecular profiles from tissues with a seconds-long turnaround time that could be inform surgical and clinical decisions without disrupting tissue analysis workflows.

## Introduction

Clinical implementation of new technologies that can augment tissue assessment is highly desirable to improve disease diagnosis and to guide clinical and surgical decision-making (1-3). Tissue evaluation is particularly critical in the surgical excision of solid cancers to assess completeness of tumor resection and surgical margin status(4). Intraoperative tissue assessment is conventionally accomplished by palpation, imaging, and histopathological analysis of frozen tissue sections prepared from an excised surgical specimen. While these procedures have vastly improved patient care over the last century (5), frozen section analysis in particular is time and labor intensive, susceptible to tissue processing artifacts that can lead to subjective and occasionally unconfirmed diagnoses, and requires specific training with a high level of anatomical pathology expertise. In breast cancer surgeries, for example, tissue evaluation and assessment of the extent of tumor involvement through frozen section analysis can be extremely challenging because breast tissues are often difficult to freeze and section. Thus, frozen sections are infrequently used, and most mastectomy and lumpectomy specimens are processed post-operatively as permanent specimens over several days. Yet, if the final surgical pathologic evaluation finds one or more margins are positive, the patient is often referred to an additional surgery for re-excision of the involved margin. For patients undergoing breast conserving surgeries, re-excision rates average around 25-30% (6, 7), leading to increased risks for patients and healthcare costs (8). Beyond oncologic surgeries, surgical procedures for excision of tissues in benign pathologies including thyroid and parathyroid diseases are often hampered by difficulties in tissue identification due to organ similarities by gross anatomy (9-12). Failed excisions can have devastating consequences for patients including the need for re-excision surgery, additional therapies, and lifelong comorbidities.

New technologies capable of providing real-time tissue assessment during surgery could greatly assist surgical decision-making, reduce operative time, and improve patient care and outcome. In particular, technologies that enable tissue analysis and identification *in vivo* prior to excision could substantially improve precision in surgical procedures while sparing unnecessary excision of functional tissues. Several imaging and molecular technologies have been developed for intraoperative use and cancer diagnosis (13-31). Fluorescence image-guided surgery, for example, has been in development for over two decades and implemented for *in vivo* tissue identification and visualization in benign and oncologic procedures (30). Optical techniques including Raman spectroscopy (21), optical coherence tomography (13), reflectance spectroscopy (14), and stimulated Raman scattering microscopy (22) have also been proposed for surgical guidance. Several mass spectrometry (MS) techniques have been explored for untargeted molecular analysis and rapid intraoperative diagnosis of tissues (15-19, 23-28, 32). The iKnife technology, for example, uses rapid evaporative ionization MS to analyze the surgical smoke generated by electrocauterization of tissues, allowing identification of normal and diseased tissues based on the detection of mass spectral profiles (29, 32). Laser ablation MS techniques have also been proposed and explored for intraoperative tissue analysis and identification through the molecular analysis of the plume formed through laser ablation process (17, 19, 23, 24, 31). We have described the development of the MasSpec Pen technology for rapid tissue analysis and cancer diagnosis based on the untargeted detection of molecular profiles characteristic of a disease state (26, 28). The MasSpec Pen operates via a liquid extraction mechanism in which a solvent droplet gently extracts molecules from a tissue for mass spectrometry (MS) analysis, without causing tissue damage. We have previously reported performance for diagnosis of multiple cancer types using *ex vivo* human tissues acquired from tissue banks as frozen specimens and analyzed in the laboratory by our research team, and demonstrated feasibility for *in vivo* tissue analysis using a murine model of human breast cancer (26, 28). More recently, we described the development of a laparoscopic MasSpec Pen device and tested its use in a robotic surgery performed on a porcine model (33). While our previous laboratory studies with human tissues and *in vivo* testing with animal models have shown feasibility of the technology for tissue analysis and diagnosis, amenability of use for molecular analysis *in vivo* and on freshly excised tissues in human surgeries operated by surgeons within the context and complexity of the surgical environment remained undetermined. Here, we describe direct molecular analysis of *in vivo* and *ex vivo* tissues performed with the MasSpec Pen technology in 100 surgical procedures over a 12-month period. The MasSpec Pen technology was translated and incorporated into the surgical workflow and used by seven different surgeons and surgical teams to perform tissue analysis during parathyroidectomy, thyroidectomy, breast, and pancreatic surgeries for various clinical indications. Collectively, the results obtained show that MasSpec Pen technology can be successfully implemented in an operating room (OR) and used by surgeons for non-destructive molecular analysis of *in vivo* and freshly excised tissues in human surgeries.

## Results

### Design and Translation of the MasSpec Pen System for Surgical Use

We designed the MasSpec Pen as a disposable handheld device integrated to a lab-built interface that is connected to a high-performance mass spectrometer for direct, real-time molecular analysis of tissues. Detailed features of the MasSpec Pen device have been previously described(28). Briefly, the MasSpec Pen device contains a biocompatible polydimethylsiloxane tip in which a reservoir is connected to three distinct conduits and two polytetrafluoroethylene (PTFE) tubing systems. Upon contact with a tissue site, a controlled volume of water is delivered through one of the PTFE tubes to the tip reservoir (2.7 mm of diameter), which is in contact and exposed to the tissue surface. The pen tip is held in contact with the tissue for a few seconds, enabling gentle and efficient extraction of molecules. The water droplet containing the extracted molecules is then transferred via a second PTFE tube to the heated inlet of the mass spectrometer for molecular analysis. In this study, a 20 µL of water droplet was used for tissue analysis with 5 seconds contact time. The third conduit was left open to air to enable balance of pressure and assist droplet transport to the mass spectrometer. All the MasSpec Pen devices used intraoperatively were fabricated using 3D printing, assembled in the research laboratory, and transferred to the hospital facilities for autoclaving before surgical use (**Fig. S1**). Sterile water and disposable sterile syringes from the OR were used in all procedures. A Q Exactive Orbitrap mass spectrometer equipped with a newly designed “dual-path” MasSpec Pen interface was installed in an OR at a hospital affiliate of Baylor College of Medicine (Houston, TX) (**Fig. 1A**). The “dual-path” MasSpec Pen interface was designed and programmed for the clinical studies with two parallel systems that allowed simultaneous channel operation under “analysis” or “priming” mode (**Fig. S2**). Under the “priming” mode, the syringe pump was activated by pressing a “refill” button to fill the PTFE tubing system with water. At the time of tissue analysis, a foot pedal was used to activate the system under the “analysis” mode and trigger the delivery of the water volume to the MasSpec Pen tip. After 5 seconds of tissue contact time, subsequent aspiration of the droplet to the mass spectrometer and a water flush of the system was performed automatically, as previously described(28). Note that this new interface was developed to improve operational efficiency and did not entail any engineering or methodological changes to the device itself. To facilitate communication of analysis status within the surgical suite, an LED system was incorporated within the MasSpec Pen interface to announce data acquisition status during surgery. A yellow light was used to indicate solvent delivery to the MasSpec Pen tip, a purple light was used to indicate the tissue contact period, and a blue light was used to indicate solvent transport from the pen to the mass spectrometer. Audio feedback, which included a simple “ping” noise, was also incorporated to provide immediate feedback on the completion of one analysis to the attending surgeon(s) so that the surgeon(s) can remove the probe from the tissue site.

**Fig. 1.**
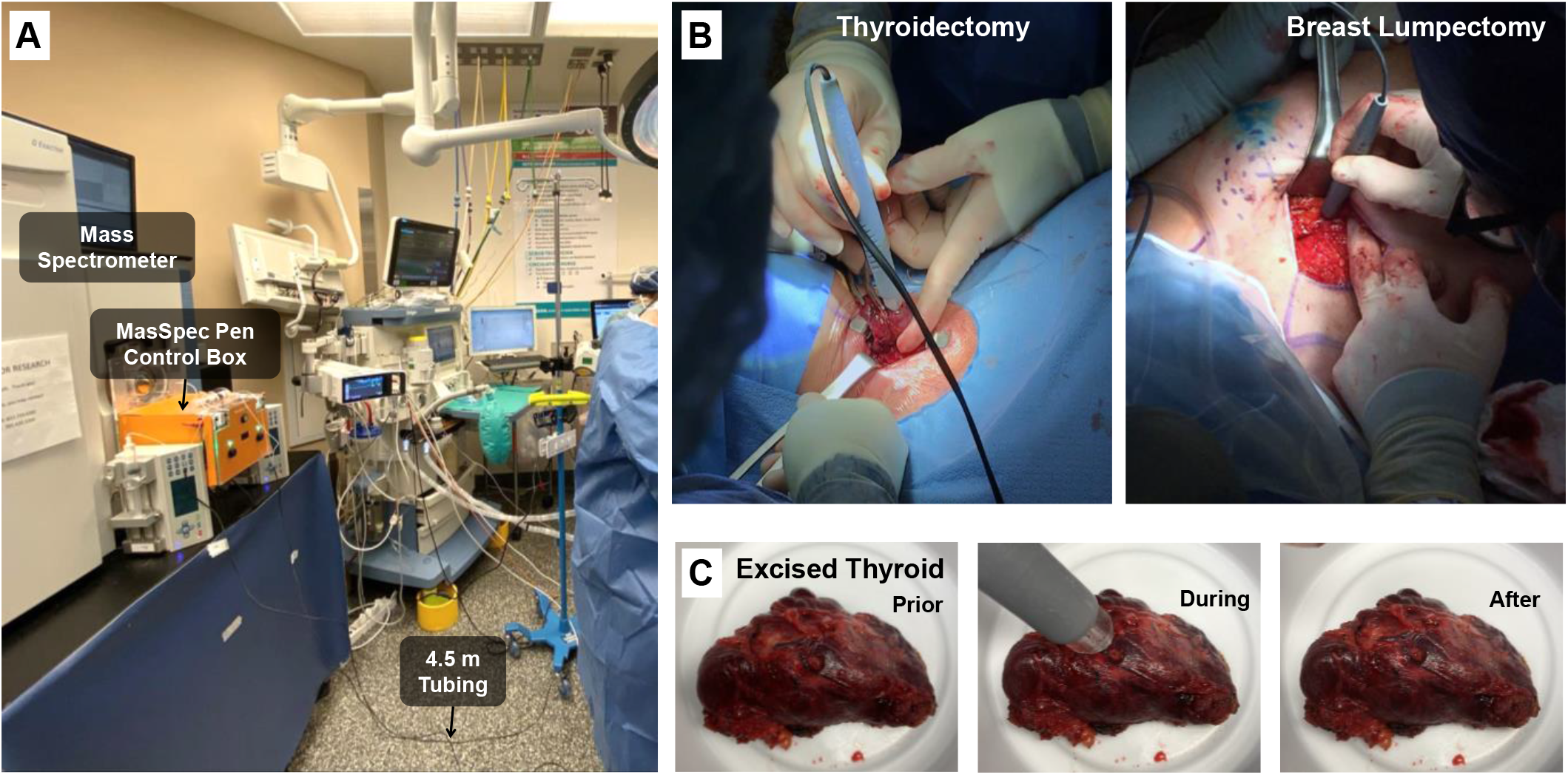
Incorporation and intraoperative use of the MasSpec Pen for *in vivo* and *ex vivo* tissue analyses in the OR. (A) A Q Exactive Orbitrap mass spectrometer equipped with a MasSpec Pen Control Box was installed in the OR. MasSpec Pen devices with 4.5 m tubing were fabricated in the laboratory and autoclaved in the hospital prior to surgical use by the surgeons. The photo was obtained during *in vivo* MasSpec Pen use in a thyroidectomy surgical procedure. (**B**) Images showing *in vivo* MasSpec Pen analysis f a thyroid gland (TH0001, left) during thyroidectomy procedure, and analysis of breast tissue in a lumpectomy prior to surgical excision (BR0006, right). (**C**) Images of a freshly excised thyroid specimen prior, during, and after *ex vivo* MasSpec Pen analysis.

The mass spectrometer was placed in the OR outside the sterile field at a ∼2 m distance from the operating table, thus requiring an extension of the tubing length beyond the 1.5 m length that we had previously used in our laboratory studies (26, 28). Different MasSpec Pen tubing lengths (1.5 m, 3.0 m and 4.5 m) were tested in the laboratory to evaluate potential impact on data quality and analysis time prior to use in the OR (**Supplementary Discussion**). As observed in the mass spectra shown in **Fig. S3**, similar mass spectra were detected for the tube lengths tested (average cosine similarity of 0.93 (n=12)), indicating that the molecular information obtained is reproducible and largely independent of tube length (**Table S1**). Additionally, droplet transfer time from the pen tip to the mass spectrometer was measured for each length tested, yielding 3.8 s ± 0.5 s (n=10), 5.8 s ± 0.7 s (n=10), and 7.5 s ± 0.4 s (n=10), for tube lengths of 1.5 m, 3.0 m, and 4.5 meters, respectively (**Table S2**). Thus, to maximize safety in our study by allowing flexibility in the placement of the tubing between the operating table and the mass spectrometer and avoid a potential tripping hazard, the 4.5-meter MasSpec Pen device was implemented in the OR, yielding a total analysis time of ∼15 seconds from tissue contact to completion of mass spectra data acquisition without impact on data quality.

### Clinical use by Surgeons and Surgical Staff

The MasSpec Pen technology was used by 7 different surgeons and surgical teams to analyze multiple tissue sites *in vivo* along with freshly excised tissue specimens during 100 surgical procedures over a 12-month period (September 2018-August 2019). Prior to each surgery, sterilized MasSpec Pen devices were brought onto the sterile field by the surgical technologists and connected to the interface and set up by the research team. To perform the analyses, the surgeon positioned the MasSpec Pen tip on the desired tissue site and the system was activated via a foot pedal. The clinical staff used the MasSpec Pen following an informal and short training (∼5 minutes) from the research team. Note that the research team was not permitted to cross onto the sterile field nor operate the devices for *in vivo* or *ex vivo* use during the surgical procedures.

**Table 1** and **Table S3** provide detailed information on patient demographics, type of surgery, clinical indication, and the analyses performed for each patient in the study. In total, 270 MasSpec Pen devices were used to collect mass spectral profiles from 715 *in vivo* and *ex vivo* analyses of healthy and diseased thyroid, parathyroid, lymph node, breast, pancreatic and bile duct tissues, with a range of 1-6 MasSpec Pen devices used per surgery. While multiple analyses were obtained with the same MasSpec Pen, a different MasSpec Pen device was used for different tissue types to prevent any potential cross-contamination between analyses (**Supplementary Discussion** and **Fig. S4**). The MasSpec Pen devices were handed to the surgeon by the surgical technologist in the same way as the other surgical tools. **Fig. 1B** shows images of the MasSpec Pen being used to analyze a thyroid on the operative field prior to its excision during a thyroidectomy (left image, TH0001) and breast cancer tissue prior to excision during a bilateral lumpectomy (right image, BR0006). A video of the MasSpec Pen being used in the thyroidectomy surgery is provided in **Movie S1**. Images of a freshly excised thyroid specimen before, during, and after MasSpec Pen analysis are shown in **Fig. 1C**. No apparent macroscopic damage to any of the tissues analyzed *in vivo* or *ex vivo* due to MasSpec Pen analysis was observed by the surgical and research team, which corroborates our previous findings (28). Accordingly, while some macroscopic blood staining was expected and noted on the tissue contact portion of the device, no observable tissue fragments were seen in the pen tip or tubing system (**Fig. S5**).

**Table 1.** Summary of patient demographics, clinical indications, and procedures.

|  | Demographics |  |  |  |  |  | Clinical Indication and Procedure |  |
| --- | --- | --- | --- | --- | --- | --- | --- | --- |
|  | Number of Patients (N) | Median Age, Years | Age Range, Years | Gender (Female, Male) | Ethnicity (Hispanic or Latino, not Hispanic or Latino) | Race (Asian, Black or African American, Native Hawaiian or Other Pacific Islander, White or Caucasian, Other) | Indication (N) | Procedure (N) |
| **Thyroidectomy** | 33 | 42 | 26 - 70 | (28, 5) | (5, 28) | (4, 8, 0, 18, 3) | Graves' Disease (6), Indeterminate Nodule (11), Papillary Thyroid Cancer (7), Toxic Multinodular Goiter (3), Other (6) | Hemithyroidectomy (16), Total Thyroidectomy (14), Other (3) |
| **Parathyroidectomy** | 42 | 59 | 31 - 81 | (32, 10) | (5, 37) | (0,10, 1, 30, 1 ) | Primary Hyperparathyroidism (35), Secondary Hyperparathyroidism (1), Tertiary Hyperparathyroidism (6) | Parathyroidectomy (22), Subtotal Parathyroidectomy (13), Revisional Parathyroidectomy (7) |
| **Breast** | 11 | 58 | 45 - 84 | (11, 0) | (3, 8) | (0, 1, 1, 9, 0) | Invasive Ductal Carcinoma (2), Infiltrating Lobular Carcinoma (4), Other (5) | Mastectomy (1), Bilateral Mastectomy (5), Lumpectomy (5) |
| **Pancreas** | 14 | 62.5 | 38 - 77 | (7, 7) | (4, 10) | (0, 1, 0, 12, 1) | Pancreatic Adenocarcinoma (4), Pancreatic Mass (5), Other (5) | Whipple (12), Distal Pancreatectomy (2) |

### Molecular Analysis of In vivo and Freshly Excised Tissues

Rich molecular profiles were detected intraoperatively from *in vivo* and *ex vivo* MasSpec Pen analyses, showing high relative abundances of various biologically relevant molecules including metabolites and lipid species previously characterized as potential disease markers in human tissues (18, 26-29, 34). **Fig. 2** shows negative ion mode mass spectra obtained from various tissue types analyzed *in vivo* during separate surgical procedures. Proposed attributions, molecular formulae, and mass errors for annotated species are provided in **Table S4**. The MasSpec Pen was used in a total of 75 endocrine surgeries, including 42 parathyroidectomies and 33 thyroidectomies. **Fig. 2A** shows an *in vivo* mass spectrum obtained from a right thyroid gland prior to its excision during a hemithyroidectomy performed on a patient with papillary thyroid cancer (TH0003). Among many peaks, high relative abundance of *m/z* 126.904 (iodine), *m/z* 175.024 (ascorbic acid), *m/z* 215.033 (hexose), *m/z* 255.234 (fatty acid [FA] 16:0), *m/z* 281.250 (FA 18:1), *m/z* 303.234 (FA 20:4), *m/z* 742.543 (glycerophosphoethanolamine [PE] 36:2), *m/z* 750.549 (PE plasmalogen [PE-P] 38:4), *m/z* 766.543 (PE 36:2), *m/z* 788.549 (glycerophosphoserine [PS] 36:1), *m/z* 822.477 (unidentified), *m/z* 835.537 (glycerophosphoinositol [PI] 34:1), and *m/z* 885.555 (PI 38:4), were detected. These molecular species were consistently detected in the mass spectra from thyroid tissues analyzed *in vivo* across various surgical procedures, as shown in the top panel **Fig. 3A** for a benign thyroid analyzed *in vivo* with the MasSpec Pen during a subtotal parathyroidectomy procedure for tertiary parathyroidism (PT0008). Similar species were also detected intraoperatively on excised thyroid tissue, as shown in **Fig. 3A** (middle panel) for the same procedure (PT0008), and have been previously described in banked thyroid tissues analyzed in the laboratory *ex vivo* using the MasSpec Pen (**Fig. 3A**, bottom panel) (28). When amenable and at the surgeon’s discretion, multiple analyses were performed with the same MasSpec Pen device on various tissue areas. For example, the mass spectra obtained from three consecutive analyses performed with the same MasSpec Pen device on an excised parathyroid during a total parathyroidectomy for primary hyperthyroidism (PT0010) is shown in **Fig. S6**. Several metabolites, including ascorbic acid and glutathione, as well as lipids species including PE, PS, and PI, were detected in the mass spectra obtained from the three analyses performed with the same device. *In vivo* analysis of the same parathyroid gland was also performed prior to its excision, yielding a comparable molecular profile with high relative abundances of the same metabolites and lipid species detected *ex vivo* (**Fig. S6**).

**Fig. 2.**
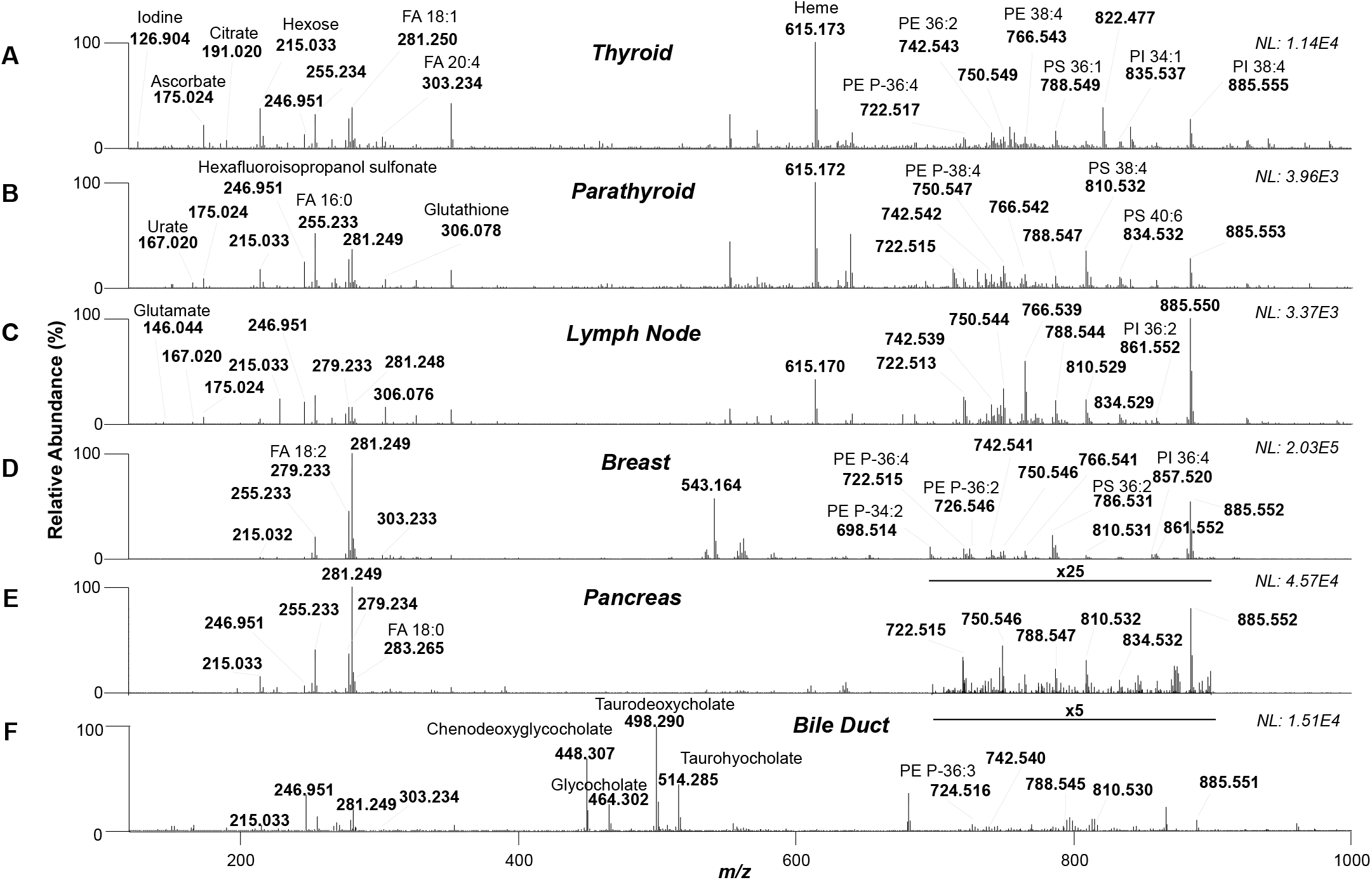
Mass spectra obtained from *in vivo* MasSpec Pen analyses of various tissues in multiple surgical procedures. Molecular data were obtained from (**A**) thyroid tissue during a right hemithyroidectomy procedure for papillary thyroid cancer (TH0003), (**B**) right inferior parathyroid during a parathyroidectomy procedure for primary hyperparathyroidism (PT0040), (**C**) lymph node during a total parathyroidectomy procedure for papillary thyroid cancer (TH0011), (**D**) breast tissue during a right mastectomy for grade III intraductal carcinoma (BR0001), (**E**) pancreatic tissue during a Whipple procedure for a pancreatic mass (PN0011), and (**F**) bile duct margin during a Whipple procedure for a cholangiocarcinoma (PN0004). Annotations for a subset of species are provided. Molecular formulas and mass errors are provided in Supplementary Table 4. Abbreviations: FA – fatty acid, PE – glycerophosphoethanolamine, PS – glycerophosphoserine, PI – glycerophosphoinositol. Total ion count (NL) for the mass spectra are shown in each panel.

**Fig. 3.**
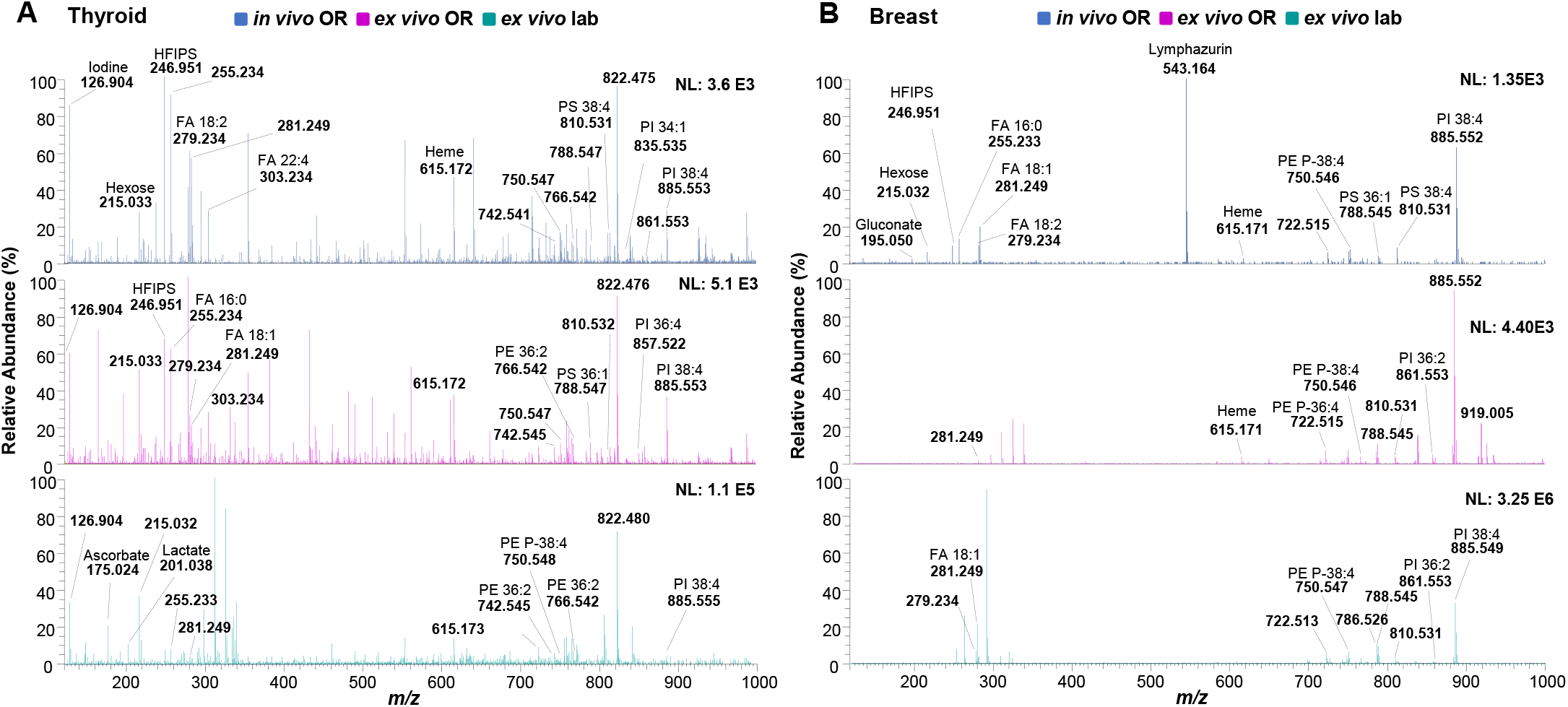
Comparison between mass spectra obtained with the MasSpec Pen *in vivo* and *ex vivo*. **(A)** Comparison between mass spectra obtained *in vivo* (average of 9 scans) and *ex vivo* (average of 11 scans) in the OR from the analysis of the same benign thyroid with the MasSpec Pen during a subtotal parathyroidectomy procedure for tertiary parathyroidism (PT0008), and *ex vivo* mass spectrum (average of 7 scans) obtained from a thawed banked benign thyroid tissue that was previously acquired in the laboratory using the same instrument(28). **(B)** Comparison between mass spectra obtained *in vivo* (average of 12 scans) and *ex vivo* (average of 12 scans) in the OR from the same breast tissue in a lumpectomy procedure (BR0006), and the *ex vivo* mass spectrum (average of 9 scans) obtained from a thawed banked benign breast tissue acquired in the laboratory using the same instrument(28). Annotations for a subset of species are provided. Total ion count (NL) for the mass spectra are shown in each panel.

*In vivo* analyses were also performed in 11 breast cancer operations (6 mastectomies and 5 lumpectomies), yielding mass spectra with high relative abundances of several metabolites and lipids previously characterized in breast tissues with the MasSpec Pen and other MS techniques (18, 28, 34). For example, **Fig. 2D** shows the mass spectrum obtained from breast tissue *in vivo* during a right mastectomy performed on a patient with grade III intraductal carcinoma (BR0001). As observed, high relative abundance of *m/z* 279.233 (FA 18:2), *m/z* 281.249 (FA 18:1), *m/z* 722.515 (PE-P 36:4), *m/z* 726.546 (PE-P 36:2), *m/z* 750.546 (PE-P 38:4), *m/z* 786.531 (PS 36:2), *m/z* 788.549 (PS 36:1), *m/z* 810.531 (PS 38:4), *m/z* 857.520 (PI 36:4), *m/z* 861.552 (PI 36:2), and *m/z* 885.552 (PI 38:4), were detected among many other peaks. A sentinel lymph node (SLN) biopsy was also performed during the procedure on patient BR0001, and the specimen was analyzed *ex vivo* using the MasSpec Pen prior to processing for frozen section analysis (**Fig. S7**). Analyses of lymph nodes both *in vivo* and *ex vivo* were conducted in 5 out of the 11 breast cancer procedures (**Table S3**). *Ex vivo* measurements of excised breast specimens were also performed in all but one of the procedures (BR0008, completion mastectomy). **Fig. 3B** shows the mass spectra obtained *in vivo* of a healthy breast tissue in a breast lumpectomy procedure prior to excision (BR0006), the mass spectra obtained *ex vivo* from the healthy breast tissue immediately following its resection, and the mass spectra obtained *ex vivo* from a banked healthy breast tissue previously analyzed in the laboratory using the MasSpec Pen(28). As observed, several metabolites, fatty acids, and lipids were commonly detected in the *in vivo* and *ex vivo* analyses. These molecular species were also detected in the mass spectra obtained from *ex vivo* MasSpec Pen analysis of an excised breast tissue during a second right breast lumpectomy for re-excision of a positive ductal carcinoma *in situ* margin (BR0003, **Fig. S8**).

The MasSpec Pen was also used to analyze pancreatic tissues *in vivo* and *ex vivo* during 14 pancreatic surgical procedures. As shown in **Fig. 2E** (PN00011), the mass spectra profiles obtained from pancreatic tissue *in vivo* presented high relative abundance of metabolites and fatty acids at the lower *m/z* range (*m/z* 200-300), including *m/z* 215.033 (hexose), *m/z* 255.233 (FA 16:0), *m/z* 279.234 (FA 18:2), *m/z* 281.249 (FA 18:1), and *m/z* 283.265 (FA 18:0), while complex lipids including *m/z* 722.515 (PE P-36:4), *m/z* 750.546 (PE P-38:4), *m/z* 810.532 (PS 38:4), *m/z* 834.532 (PS 40:6), and *m/z* 885.552 (PI 38:4) were also detected in the higher *m/z* range, although at lower relative abundances. Many of these molecular species have been previously detected in human pancreatic tissue sections using MS imaging (27). Analysis of excised pancreatic tissues were also performed in 10 of the pancreatic procedures. **Fig. S9** shows the molecular profiles from the *ex vivo* analysis of pancreatic tissue performed during a Whipple procedure for a pancreatic mass (PN0006), in which many of the molecular species detected *in vivo* were also observed. Further, analysis of bile duct tissues was performed in two of the Whipple procedures. *In vivo* analysis of the bile duct margin during a Whipple procedure for a cholangiocarcinoma (PN0004) yielded a distinct mass spectrum with high relative abundance of bile acids in addition to metabolites and lipid species (**Fig. 2F**). For example, ions at *m/z* 448.307, *m/z* 464.302, *m/z* 498.290, and *m/z* 514.285 were identified as the bile acids chenodeoxyglycocholate, glycocholate, taurodeoxycholate, taurohyocholate, respectively (35). Collectively, these results demonstrate that the MasSpec Pen technology allowed surgeons and the surgical team to effectively acquire rich molecular data directly from tissues in real time during surgical procedures.

### Intraoperative Detection of Blood and Exogeneous Species

The presence of residual blood at the tissue surface *in vivo* and on the freshly excised tissues was presumed to be a source of potential interference in intraoperative molecular analysis using the MasSpec Pen. As anticipated, detection of blood-related ions was observed at generally higher relative abundances in the mass spectra collected from *in vivo* and freshly excised tissues when compared to *ex vivo* mass spectra previously acquired in the laboratory with banked tissues (**Fig. 3 and Fig. S10**). In particular, *m/z* 615.171, identified as deprotonated heme (**Fig. S10 and Fig. S11**), and a series of multiply charged ions in the *m/z* 1200-1800 range, tentatively identified as deprotonated hemoglobin subunits (**Fig. S12**), were detected in the majority of the mass spectra acquired intraoperatively, at varied relative abundances. Heme and hemoglobin were also observed in the mass spectra obtained when directly analyzing human blood in the laboratory using the MasSpec Pen (**Fig. S13**), with heme at *m/z* 615.171 detected at high relative abundance. In addition to blood-related molecules, a series of sodium chloride cluster peaks from *m/z* 200-1300 associated to the saline solution used for tissue irrigation were also observed in the mass spectra obtained in several surgical procedures (**Fig. S14**). While the high mass resolution provided by the Orbitrap mass spectrometer allowed metabolites and lipid ions to be resolved and detected among the saline-related peaks, the mass spectra was largely convoluted and thus sterile water was used as a preferred irrigation solution by the surgical team during MasSpec Pen experiments. Interestingly, detection of a compound at *m/z* 246.951 was consistently observed in the mass spectra acquired *in vivo* and in freshly excised tissues intraoperatively from patients (**Fig. 2, 4** and **5A**). This species had been previously described in *ex vivo* tissues as a heavy fluorinated compound (C_3_HF_6_SO_4_^-^) of possible anesthetic origin (36). The detection of *m/z* 246.951 in the mass spectra collected intraoperatively provided evidence of the identification of this species as hexafluoroisopropyl sulfonate (HFIPS), a suspected secondary metabolite of sevoflurane, the anesthetic compound used during the surgical procedures. To corroborate this hypothesis, we synthesized HFIPS and performed tandem MS experiments (**Supplementary Information**) (37). The fragmentation pattern and product ions of the synthesized compound at *m/z* 246.950 were consistent with those obtained from *m/z* 246.951 detected from tissues using the MasSpec Pen, thus supporting its identification as HFIPS (**Fig. 5C** and **Fig. S15**). Moreover, an ion at *m/z* 543.164 was detected *in vivo* and *ex vivo* in breast and lymph node tissues analyzed in breast surgeries (**Fig. 2D** and **Fig. 3B**). This ion was identified as isosulfan blue (lymphazurin), a blue dye used in breast cancer procedures for as a SLN mapping agent(38), which could be visibly observed in the breast tissues.

## Discussion

In this study, we report the clinical implementation of the MasSpec Pen technology for molecular analysis of tissues during human surgeries. The MasSpec Pen technology was used directly by surgeons to effectively and rapidly acquire molecular data in real time during surgical procedures. To facilitate clinical use by surgeons and clinical staff in the OR, the MasSpec Pen interface was re-designed to incorporate audio and visual feedback, thus improving communication of the analysis status to the clinical staff with minimal disruptions to the surgical workflow. Further, the interface was programmed with two parallel systems to facilitate use of a new device when desirable to the surgeon, allowing negligible time lag when using a new device and facilitating use in the OR. Similar to other instrumentation routinely used in the OR, the sterilized devices were handled by surgical technologist to the surgeon and replaced as desired, with a range of 1-6 devices used per surgery. To maximize safety for patients and surgical staff, a MasSpec Pen tube length of 4.5 meters was used in all surgical procedures. While shorter tube lengths could have been employed, the 4.5 meters tube yielded data quality similar to what obtained with shorter tubing lengths at a slightly longer droplet transfer time and was thus implemented to facilitate maneuverability within the OR. To minimize risks of infection or other potential side effects to the patients, all materials were sterilized by autoclaving before use, and sterile water and sterile syringes were used. The surgical teams indicated the MasSpec Pen analysis procedure was efficient, well-tolerated, and no device specific intraoperative or postoperative complications to patients were reported.

The MasSpec Pen allowed gentle and effective extraction of biomolecules *in vivo* and from freshly excised tissues using a small volume (20 µL) of sterile water, without causing apparent macroscopic tissue damage. In total, mass spectral profiles were obtained from 715 *in vivo* and *ex vivo* analyses of a total of 177 organs during 100 surgeries, including healthy and diseased thyroid, parathyroid, lymph node, breast, pancreatic and bile duct tissues. The capability to obtain molecular information *in vivo* and from fresh specimens while preserving tissue integrity is critical to ensure patient safety and avoid damage to healthy tissues, as well as paramount to facilitate integration with standard-of-care clinical procedures. Other MS technologies proposed for intraoperative tissue analysis rely on more invasive methods to achieve molecular analysis, such as tissue vaporization or laser ablation, which may impede pathologic evaluation of the tissue area analyzed and/or damage tissues *in vivo*. In previous studies, we have shown compatibility of the MasSpec Pen analysis with pathology, and have used common surgical inks to mark the tissue area analyzed for subsequent staining and evaluation (26). In this study, no limitations to histologic tissue processing or diagnosis were identified by the pathology department due to MasSpec Pen use in any of the surgical procedure and tissues analyzed. These observations strengthen the potential of the MasSpec Pen for intraoperative use and incorporation into clinical workflows.

The molecular profiles obtained intraoperatively from *in vivo* and *ex vivo* MasSpec Pen analyses showed high relative abundances of various biologically relevant molecules including metabolites and lipid species previously characterized as disease markers in human tissues using the MasSpec Pen and many other MS techniques (18, 26-29, 34). Several of the detected molecules have been described in different studies to have biological significance in disease progression or treatment, such as glutamic acid (39), ascorbic acid (40), and glutathione (41). As observed in the data obtained for several tissues and surgeries, the mass spectra acquired from the same tissue analyzed *in vivo* and *ex vivo* immediately following its resection presented high relative abundance of similar molecular ions, providing evidence that the gentle liquid extraction process employed by the MasSpec Pen allows molecular analysis in both living and freshly excised tissues. As expected for tissues analyzed in the surgical procedures, detection of blood-related ions was observed in the mass spectra obtained from *in vivo* and freshly excised tissues (**Figs. 3, 4, S7, S8, S10, S11**, and **S12**). Spectral interferences due to blood during intraoperative tissue analysis have been previously described for other technologies. For example, presence of blood has been shown to affect the fluorescence of molecular probes used in fluorescence-guided brain tumor surgeries, causing both quenching and increases in overall signal (42). Moreover, contamination of optical devices with blood have been shown to block light transmission in fluorescence-guided procedures (42). These challenges have been largely mitigated though the implementation of device cleaning protocols and development of computational methods for spectral subtraction of the blood-related effects observed in the fluorescence signal (43). Using MS, the most prominent spectral features associated with blood can be identified with high mass accuracy measurements, high mass resolution, and the chemical specificity provided by MS. In the intraoperative data acquired in our study, heme and hemoglobin ions were detected at varied relative abundances in the mass spectra obtained. For example, a low relative abundance of heme (∼2%) was observed in the *in vivo* mass spectra acquired from breast tissue (**Fig. 2D**), while a higher relative abundance of various fatty acids and lipid species, such as FA(18:1) at *m/z* 281.249 (100%) and PI(38:4) at *m/z* 885.544 (∼50%), was observed. In the *in vivo* mass spectra acquired from a normal parathyroid tissue (**Fig. 2A**), metabolites and lipids species were detected at varied relative abundance while heme was detected as the highest intensity peak (100%) in the mass spectra. Importantly, metabolite and lipid species were not observed at high relative abundances in the mass spectra acquired from blood alone (**Figure S13**), suggesting that the presence of residual blood on *in vivo* and freshly excised tissues does not strongly contribute to or preclude detection of lipid and metabolite profiles from tissues. However, additional studies are needed to systematically evaluate the effect of blood and other potentially interfering compounds on the molecular profiles obtained from tissues. Other exogenous compounds, such as HFIPS, a product of the anesthetic compound used in surgical procedures, and isosulfan blue, a dye used for SLN mapping in breast surgeries, were also detected in various mass spectra acquired without hindering detection of endogenous molecular species. Collectively, the intraoperative data obtained shows evidence that the MasSpec Pen allows extraction and detection of biologically relevant molecules *in vivo* and from freshly excised specimens, supporting the continued development of this technology for clinical use and tissue identification.

A few operational and technical challenges were identified during the intraoperative studies. Most notably, a decrease in instrument performance due to contamination of the first-stage ion optics following consecutive instrument use during multiple surgeries was noted within the first two months of the pilot clinical study, similar to what reported by others (20, 32). Moreover, a momentary loss in system status occurred in 9 of the 100 surgeries due to vacuum instability, requiring instrument reboot and triggering an increase in instrument pressure. In one of the 100 surgeries (PN0010), the instrument performance was observed to be below operational standards, precluding data acquisition. Following this occurrence, we implemented standard procedures for cleaning the instrument ion optics and calibration at approximately every 10 surgeries during subsequent months. Multiple practical approaches to refine system design are currently being explored to reduce contamination and improve instrument robustness and ease of maintenance for surgical use. For example, a rapid and automated washing flush that is diverted from the mass spectrometer into a waste bottle is currently being incorporated into the programmed analysis steps to reduce buildup in the ion optics and the total volume of water introduced to the mass spectrometer per analysis, as well as to improve reusability of the device during surgery. Efforts to reduce contamination of the mass spectrometer during *in vivo* use by redesign of the ion optics components have been reported by other methods (20, 32), and may be explored in future iterations of a clinical mass spectrometer to improve robustness and ease of maintenance. Concerns regarding the Orbitrap mass spectrometer size (95 x 83 x 91 cm) and vacuum pump noise levels (∼70 dB) were reported by OR personnel. However, no identified effects on patient care or progress of the surgical procedures was reported as a result of the instrumentation. The challenges and opportunities identified in this clinical study give rise to the potential for the development of a console-type mass spectrometer as a portable wheeled unit with a footprint similar to standard OR carts to facilitate clinical implementation of the MasSpec Pen technology and allow use of a single unit across multiple ORs. We have previously tested the performance of the MasSpec Pen to differentiate normal from cancerous ovarian tissues *ex vivo* using a lower performance ion trap mass spectrometer, which is smaller and less costly than the Orbitrap, as a potential alternative mass spectrometer for selected intraoperative applications (26). Importantly, the surgical and clinical staff universally confirmed the potential of the technology warrants further investigation and development in order to benefit future patient care.

Note that there are several limitations to this study. Foremost, this is a prospective cohort pilot study performed at a single clinical center to evaluate technical feasibility of a new intraoperative technology for molecular analysis of human tissues and was not specifically powered or designed to evaluate diagnostic performance or differences in clinical outcomes. Surgical procedures for various clinical indications, as showed in **Table 1** and **Table S3**, were included in this study to reflect the variety of surgeries performed at the clinical center, entailing different surgical instrumentations, surgical techniques, as well as different clinical teams with varied specialties. Future research studies with larger cohorts of patients powered for specific types of operations and tissues, including laparoscopic and robotic procedures, are needed to validate the findings. Further, sample marking was not incorporated at this initial phase for excised specimens to avoid interfering with pathologic procedures used during frozen section analysis. A clinical protocol for tissue annotation is currently being optimized to allow precise pathological evaluation of the tissue area analyzed without interfering with current clinical workflow. Importantly, a larger cohort of patients and procedures is needed to properly evaluate the diagnostic performance of the MasSpec Pen for tissue assessment during surgery. Statistical modeling and prediction of a larger dataset will be systematically performed for each indication in consecutive studies to evaluate the diagnostic and intraoperative performance of the MasSpec Pen for tissue identification, diagnosis, and surgical margin evaluation.

To conclude, we describe the successful translation and intraoperative use of the MasSpec Pen technology for direct and rapid analysis of *in vivo* tissues on diverse operative fields and of freshly excised tissues from 100 patients undergoing a variety of surgical procedures. This study effectively demonstrates feasibility for intraoperative use of mass spectrometry by surgical teams and integration of the technology into a clinical workflow. Notably, the MasSpec Pen allowed detection of molecular profiles from tissues intraoperatively with a seconds-long analysis turnaround time that could be valuable to inform surgical and clinical decision-making. Importantly, this study showed that implementation of the MasSpec Pen technology did not impact pathologic evaluation of tissues nor cause any reported complications to the patients enrolled. Following clinical research studies will be focused on continuous intraoperative tissue analysis in larger cohorts of patients undergoing cervical, breast, and pancreatic cancer operations to evaluate diagnostic performance and potential in guiding surgical decision-making.

## Materials and Methods

### Chemicals and Reagents

Sterile water (Deerfield, Illinois, USA) commonly used during surgery was provided by the clinical staff and used for all analyses. Human whole blood was purchased from BioIVT (Westbury, NY, USA). Silicon tubing, PTFE tubing, and heat shrink tubing were purchased from Kinesis (Vernon Hills, IL, USA), MilliporeSigma (Burlington, MA, USA), and Newark element14 (Chicago, IL, USA). Luer-locks were purchased from Cole-Parmer (Vernon Hills, Illinois, USA). The other materials used to 3D-print the MasSpec Pen devices were described in detail in our previous report.(28)

### MasSpec Pen Device and Operation

The MasSpec Pen devices were produced in the laboratory as previously described (28). All devices were discarded after each surgery observing universal precaution measures.

### Dual Path MasSpec Pen Control Box

The MasSpec Pen interface/control box was designed and fabricated by the Instrument Design and Repair Services at the University of Texas at Austin Department of Chemistry. The interface houses an Arduino board (ELEGOO MEGA 2560 R3 Board), a lab-built circuit board, two three-way solenoid pinch valves (Masterflex, Gelsenkirchen, Germany). The front panel of the MasSpec Pen interface/control box is shown in **Fig. S3**. A lab-built extended metal capillary was inserted into the mass spectrometer inlet, and to the third PTFE conduit of the MasSpec Pen connected to the metal capillary through a silicon tubing. Two syringe pumps (Chemyx Inc, Stafford, TX, USA) were connected to the control box, enabling parallel analysis and priming of the MasSpec Pen devices. The control box was equipped with a switch to alternate the “priming” and “analysis” functions between the two syringe pumps. Under “priming” function mode, the syringe pump was activated by pressing the “Refill” button, allowing pumping of 2.5 mL of water to fill the 4.5 meter PTFE tube (∼ 15 seconds). At the time of tissue analysis, a foot pedal was used to activate the analysis by triggering the delivery of a discrete water volume (20 µL) to the MasSpec Pen tip, followed by 5 seconds of tissue analysis time, subsequent aspiration of the droplets to the mass spectrometer, and a water flush of the system (106 µL), as previously described(28). An LED and audio system were also incorporated in the MasSpec Pen control box interface. Note that sterile water and disposable sterile syringes were used in all procedures.

### Mass spectrometry analysis

Experiments were performed on a Q Exactive™ Hybrid Quadrupole-Orbitrap™ mass spectrometer (Thermo Fisher Scientific, San Jose, CA, USA). The mass spectrometer was installed in the operating room using a 220 Volt supply. Analysis were carried out in full scan mode in the negative ion mode at the range of *m/z* 120 to 1800, using a resolving power of 140,000, capillary temperature of 400 °C, and an S-lens radio frequency level of 100. Note that all the mass spectra data were acquired in centroid mode. High mass resolution/accuracy measurements and tandem MS analyses were performed for ion identification. Due to the strict time constraint for data acquisition in the OR, tandem MS experiments were performed in the laboratory with *ex vivo* tissues using a Q Exactive™ HF Hybrid Quadrupole-Orbitrap™ mass spectrometer (Thermo Fisher Scientific, San Jose, CA, USA). Analysis of human blood was also performed in the laboratory by depositing 10 µL of human whole blood onto a glass slide and analyzing the blood with the MasSpec Pen using the same experimental conditions used for OR tissue analysis.

### Data Analysis

Mass spectral data was analyzed using XCalibur 3.1 Qual Browser. For cosine similarity analyses of the data collected in the laboratory for different MasSpec Pen tubing lengths, four mass spectra for each analysis were first normalized to their total ion current (TIC) and then averaged. Cosine similarity analyses were performed in the CRAN R language using average mass spectra for each analysis.

### Permissions

The study was reviewed and approved by the Institutional Review Board (IRB) of Baylor College of Medicine and the IRB of the University of Texas at Austin. The intraoperative use of the MasSpec Pen was restricted to research purposes on patients pre-scheduled to undergo surgery independently of our study. Patients provided written consent to participate in the study under the approved IRB protocol. The data from the analyses were not communicated to the surgical team nor used in clinical decision-making. Patients scheduled to undergo parathyroidectomy, thyroidectomy, breast, and pancreatic cancer surgeries were approached by the clinical research assistants prior to surgery to participate in the study. Written consent was obtained for all patients that agreed to participate in the study. The research team was notified by clinical research assistants if the patient provided written consent to participate in the study so that MasSpec Pen analyses could take place during the surgical procedure. Patient information was de-identified by the clinical research assistants using a coding system, according to approved IRB protocol.

## Data Availability

Data will be available upon peer-reviewed publication of the manuscript.

## Conflicts of interest

J.Z., R.J.D., J.Q.L., C.L.F., N.K., T.E.M., W.Y., J.S., and L.S.E. are inventors in US Patent No. 10,643,832 and/or in other patent applications related to the MasSpec Pen Technology licensed by the University of Texas to MS Pen Technologies, Inc. J.Z., T.E.M., J.S., and L.S.E. are shareholders in MS Pen Technologies, Inc. T.E.M., J.S., L.S.E., and C.L.F. serve as a technology adviser, chief medical officer, chief scientific officer, and a consultant, respectively, for MS Pen Technologies, Inc.. All other authors declare no competing interests.

## Acknowledgements

This work was supported by the National Cancer Institute of the National Institutes of Health under award numbers R00CA190783 and 1R33CA229068-01A1, and by the Gordon and Betty Moore Foundation through grant GBMF8049. The authors would like to thank all the patients in the study, the surgical technologists Stephanie R. Orenstein and Corey Jones Utley, and the clinical staff at the hospital affiliate of Baylor College of Medicine. The authors would also like to thank Tim Hooper for building the MasSpec Pen interface, and Noah Giese for assistance with laboratory experiments.

## Supplementary Information

### Supplementary Discussion

#### MasSpec Pen Tube Length

Due to the distance between the mass spectrometer and the operating table in the OR (∼2m) and the safety constrains with the tubing system as a potential tripping hazard, 4.5 meters of tube length was used in the OR experiments, which is 3 times longer than what we had previously reported.(1) As such, different MasSpec Pen tube lengths were tested in the laboratory to evaluate potential impact on data quality and analysis time prior to use in the operating room. **Fig. S2** shows the mass spectra obtained from serial sections of mouse brain tissue analyzed with a 2.7 mm MasSpec Pen probe connected to polytetrafluoroethylene (PTFE) tubing with 1.5 m, 3.0 m and 4.5 m lengths. The tubing length was measured from the MasSpec Pen handheld device to the mass spectrometer inlet. Similar molecular patterns were observed in the recorded mass spectra, displaying high relative abundances of a variety of negatively charged ions, identified as lipid species typically observed from mouse brain tissue sections using MasSpec Pen and other ambient ionization techniques.(1, 2) For example, *m/z* 834.529 (identified as [PS(40:6)-H]^-^), *m/z* 885.550 (identified as [PI(38:4)-H]^-^), and *m/z* 790.539 (identified as [PE(40:6)-H]^-^)), were observed at high relative abundances in the mass spectra obtained from the grey matter of mouse brain. Please note that less similarity was observed in the mass spectra profiles in the *m/z* 120-400 range, where fluctuations in the intensities of background ions including *m/z* 297.153, *m/z* 311.168, and *m/z* 325.164 are often observed in our MasSpec Pen analyses (**Fig. S2**). These ions are not biologically related and likely arise from impurities in the water used or the PTFE tubing. Biologically related ions at the lower *m/z* range including *m/z* 146.044 (identified as glutamate), *m/z* 175.023 (identified as ascorbate), and *m/z* 255.233 (identified as FA 16:0), are consistently detected in the mass spectra. An average cosine similarity of 0.93 (n=12) was achieved for the mass spectra obtained with various tube lengths, which indicates that the molecular information obtained is reproducible and independent of tube length (**Table S1**). Additionally, the transfer time of the entire droplet volume plus flush used from the pen tip to the mass spectrometer inlet was measured for each length tested, yielding 3.8 s ± 0.5 s (n=10), 5.8 s ± 0.7 s (n=10), and 7.5 s ± 0.4 s (n=10), for tube lengths of 1.5 m, 3.0 m, and 4.5 meters, respectively (**Table S2**).

#### Contamination between MasSpec Pen Devices

In our effort to be abundantly cautious in preventing potential cross-contamination between analyses and tissue sites, different MasSpec Pen devices were used to analyze different organs or tissue sites during a surgical procedure. A “water wash” was employed between devices when surgical timing allowed to do so, in order to evaluate background signal and thus potential contamination of the system and mass spectrometer interface. The “water wash” background was collected by triggering the pedal and conducting the analysis without touching a tissue. **Fig. S4** shows the mass spectra obtained from a water background and tissue analyses during a total parathyroidectomy procedure. In the surgery, a first pen was used to analyze parathyroid tissue. After analysis, a new pen was activated and a water wash background was collected, as shown in panel A. As observed, the water background mass spectra did not present the characteristic lipid and metabolite profiles obtained from tissue analysis, regardless of previous tissue analyses performed with other MasSpec Pen devices during the surgery. Following the water wash, analyses of a lymph node were performed, yielding rich mass spectral profiles with many biologically relevant species (panel B). A third pen was then activated in the system for subsequent analysis of thyroid tissue. As seen in panel C, a background mass spectrum obtained with the third pen did not present biological peaks associated with tissue analyses. Note that variation in the mass spectra obtained from water backgrounds was commonly observed in our experiments, which we have presumably associated with variations in residual compounds from the autoclaving process, pen materials, and/or water impurities.

#### Detection of Exogeneous Species

Detection of ion species associated with two exogenous sources was observed in the data obtained intraoperatively: saline and anesthetic-related molecules. The saline solution used for tissue irrigation during surgery yielded a series of sodium chloride cluster peaks from *m/z* 200-1300 (**Fig. S14**). While metabolites and lipid ions were still detected among the saline-related peaks, sterile water was used as a preferred irrigation solution by the surgical team when possible. Interestingly, a peak at *m/z* 246.951 was observed in the mass spectra acquired from several patients. This species had also been previously detected in our laboratory by desorption electrospray ionization mass spectrometry analysis of prospectively collected specimens from endometriosis surgeries(3), and was identified as a heavy fluorinated compound (C_3_HF_6_SO_4_) of possible anesthetic origin. The detection of *m/z* 246.951 at high relative abundances in the mass spectra collected in this study with the MasSpec Pen from *in vivo* and freshly excised specimens during surgery supported the identification of this species as hexafluoroisopropyl sulfonate (HFIPS), a suspected secondary metabolite of sevoflurane, an anesthetic compound used during surgery. To corroborate this hypothesis, we performed an experiment to biosynthesize HFIPS by incubating HFIP, a known metabolite of sevoflurane produced by Cytochrome P450 2E1, with rat liver S9 and 3’-Phosphoadenosine-5’-phosphosulfate.(4) The resulting product was analyzed by ESI-MS/MS revealing similar patterns to those obtained from tissue samples, thus supporting its identification as HFIP-sulfonate (**Fig. S15**).

## Supplementary Figures

**Figure S1.**
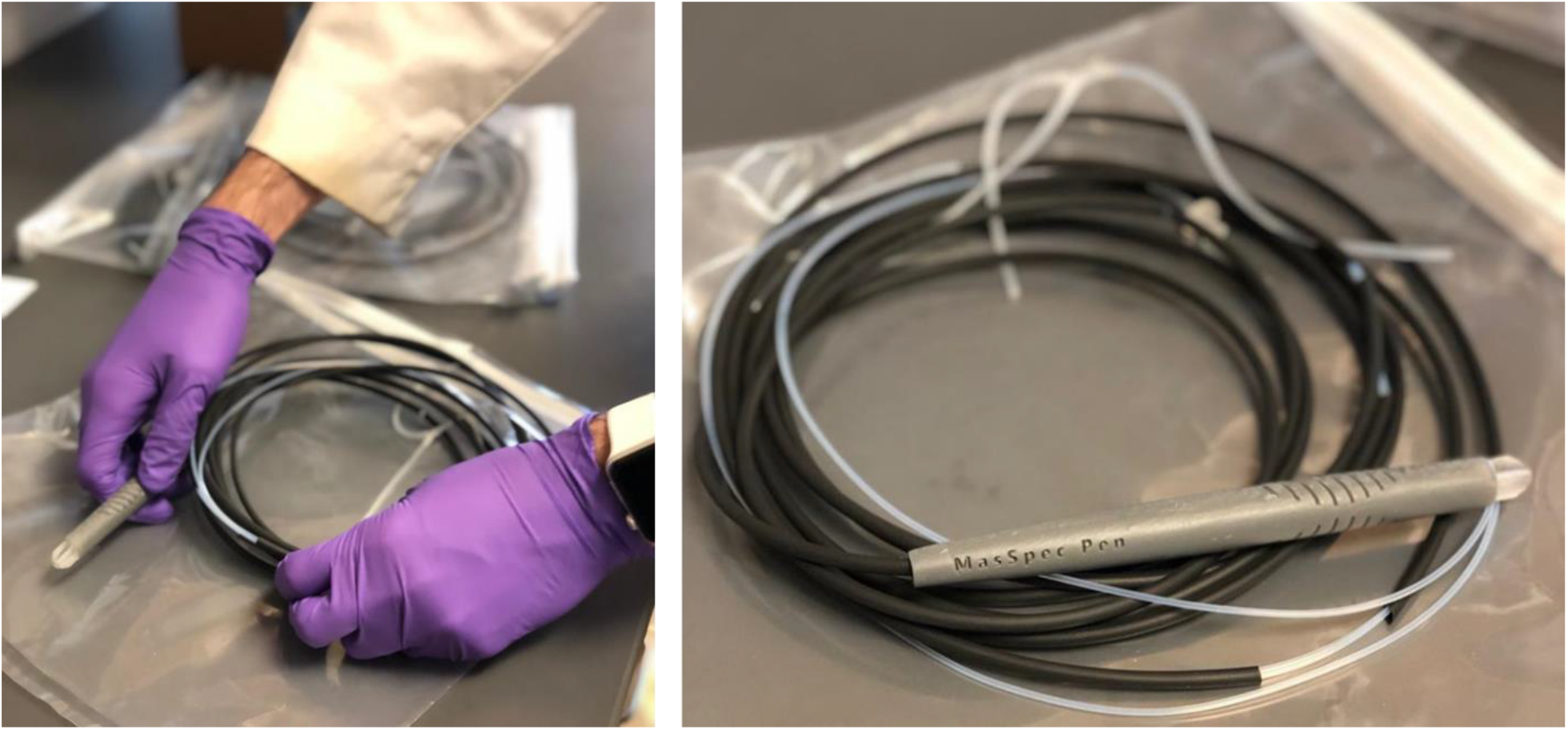
Assembling of MasSpec Pen devices. All the devices used intraoperatively were 3D printed and assembled in the laboratory at UT Austin. After assembly, each individual device was placed in sterile bag and sent to the hospital for autoclaving prior to surgical use.

**Figure S2.**
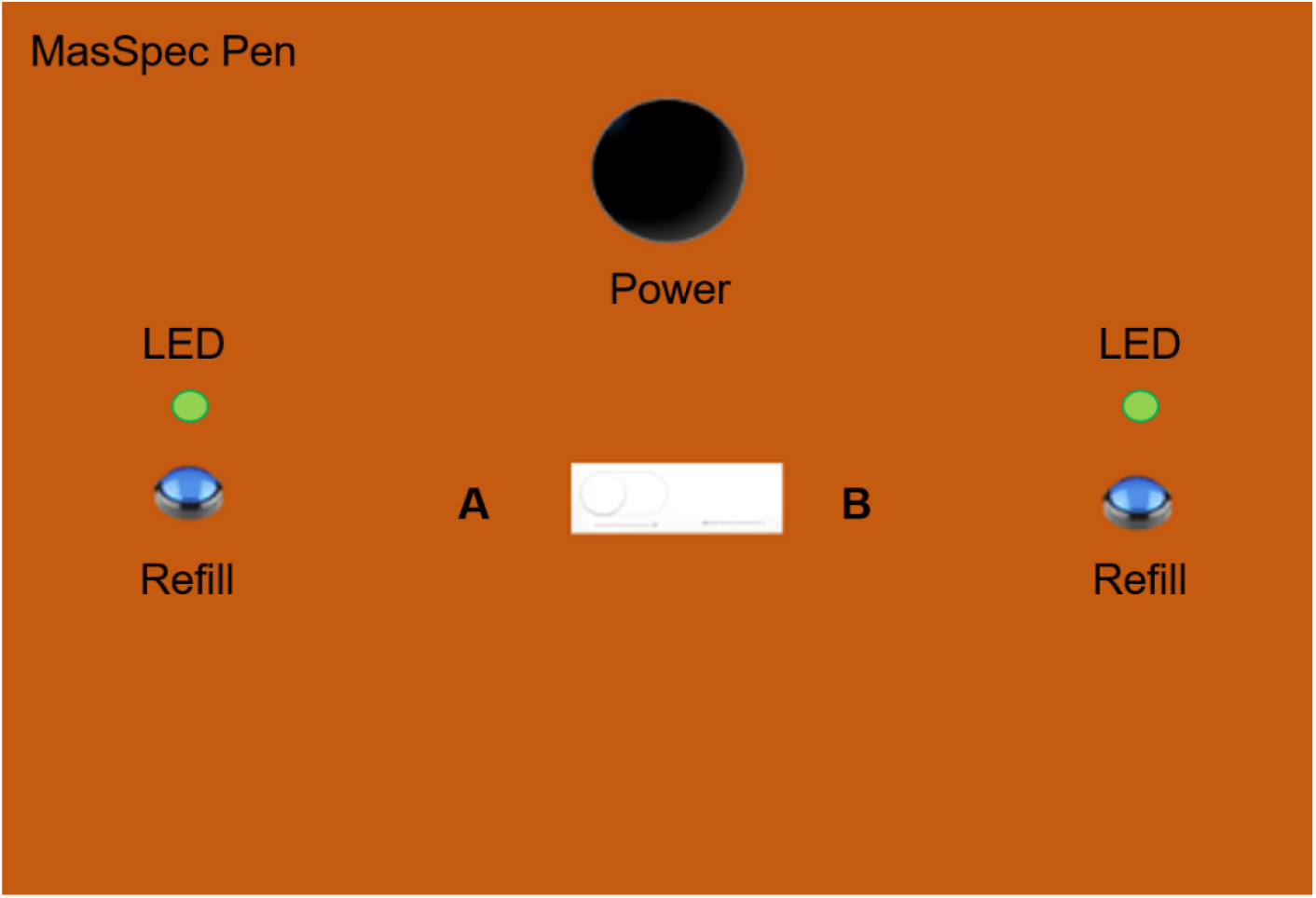
MasSpec Pen control interface front panel. The control box is composed of four buttons which are power, A&B switch, and refill. LED lights were incorporated in the panel to indicate the status of “analysis” or “prime”.

**Figure S3.**
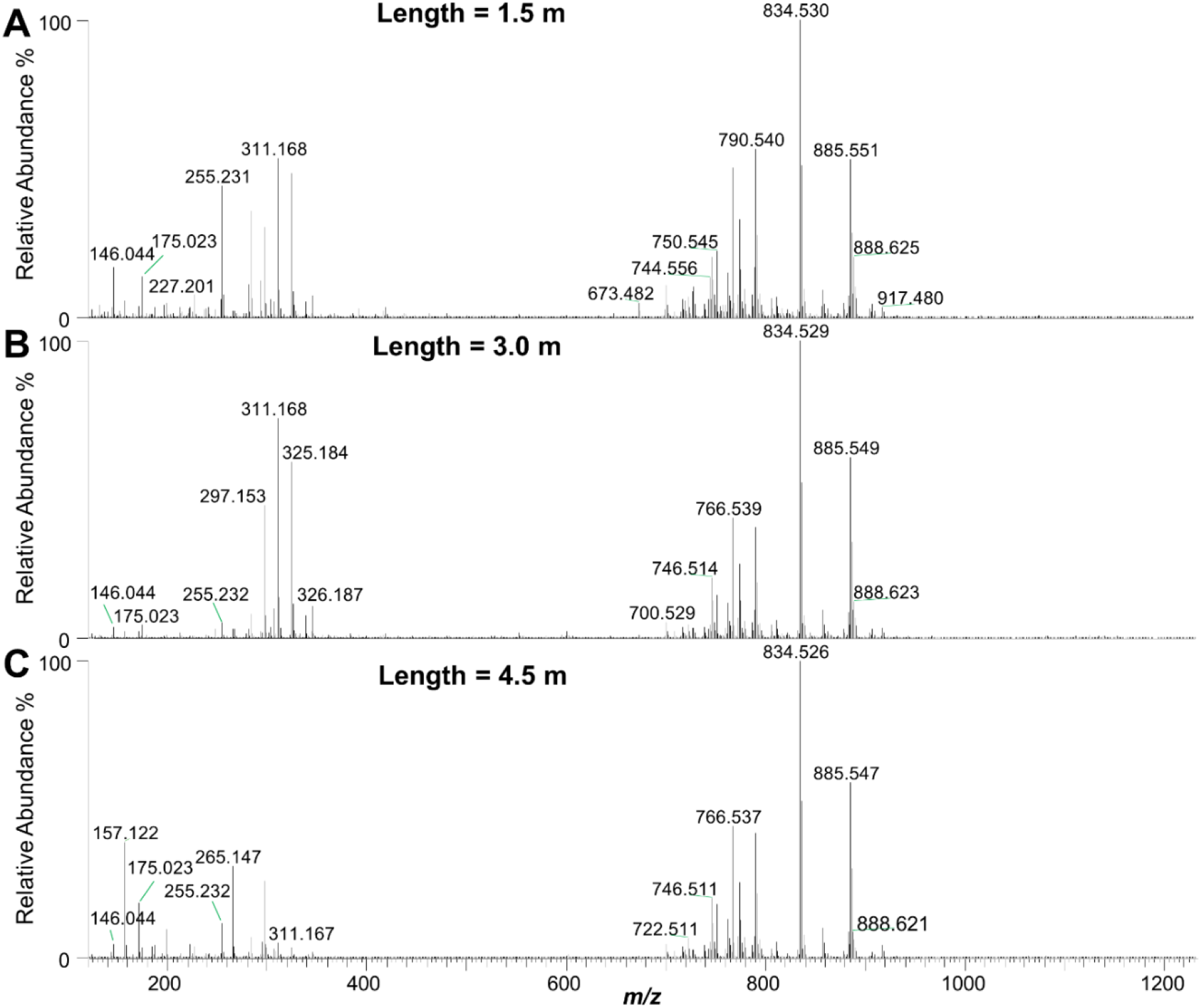
Mass spectra acquired with different MasSpec Pen tubing lengths. The data quality obtained at different tubing was evaluated using mouse brain tissue sections. As observed, similar molecular profiles were obtained at different transfer times using (A) 1.5 m, (B) 3.0 m, and (C) 4.5 m tube lengths.

**Figure S4.**
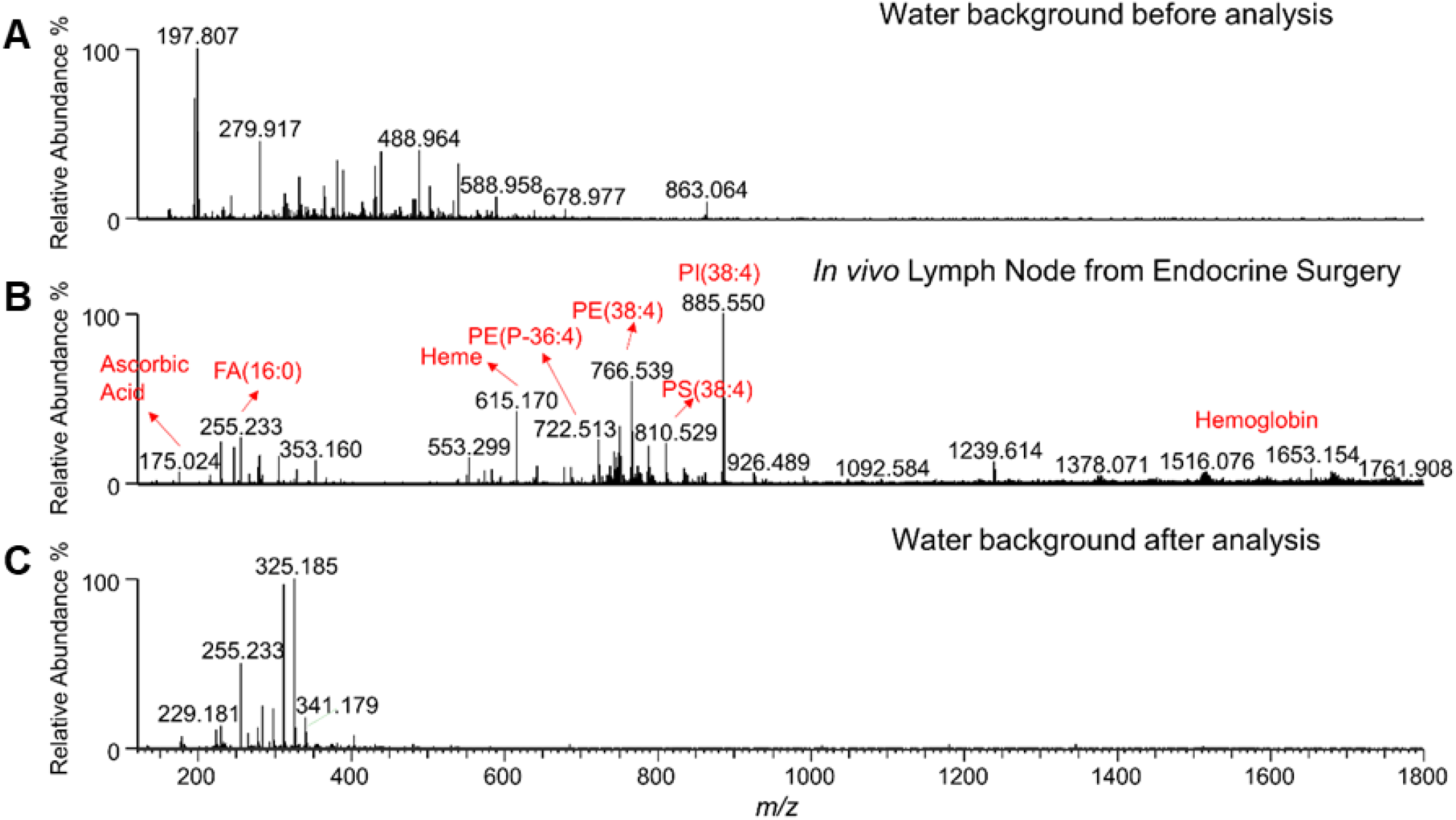
Mass spectra obtained during a total thyroidectomy for papillary thyroid cancer (TH0011). A water background mass spectrum was first collected using a new MasSpec Pen device (A) prior to sampling a lymph node in vivo (B). After tissue analysis, the MasSpec Pen was discarded, and a new device was used for subsequent sampling. (C) Water background mass spectrum obtained with the new MasSpec Pen device.

**Figure S5.**
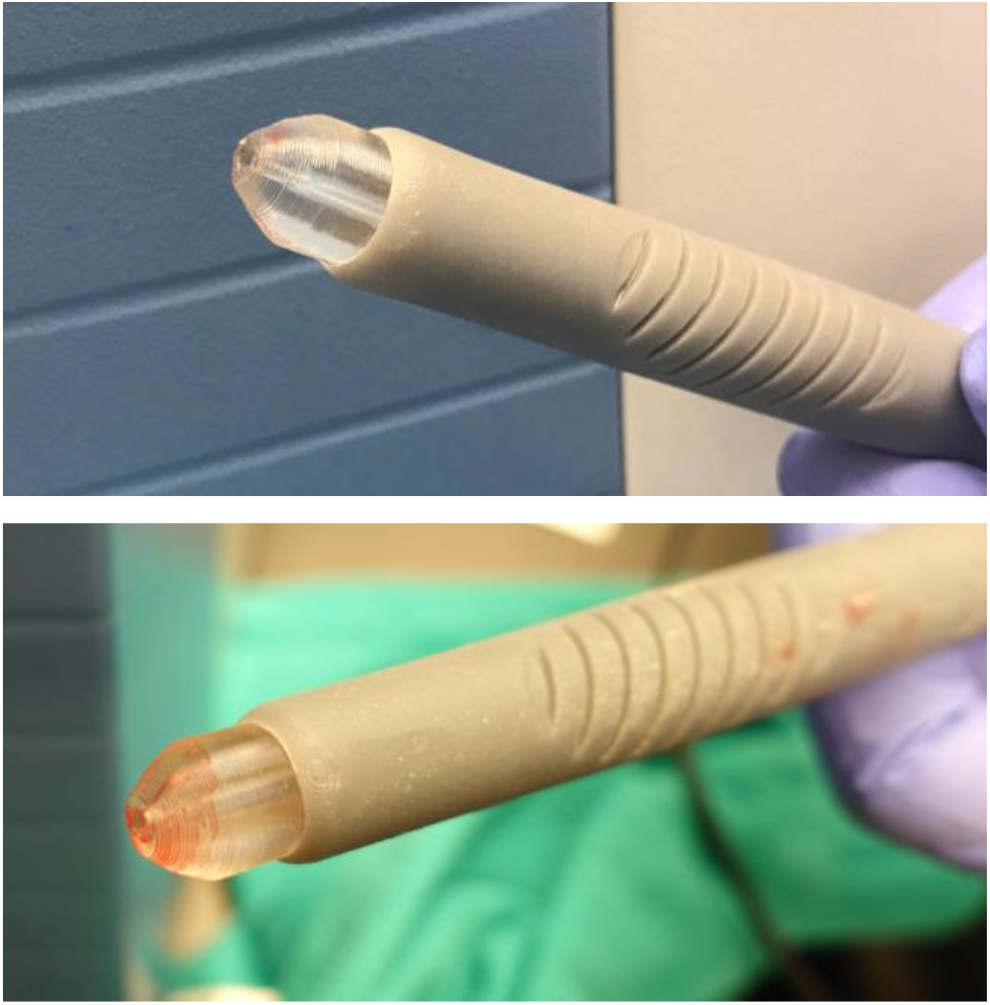
Images of two different MasSpec Pen devices after being used *in vivo* by surgeons for tissue analyses. As observed in the images, while macroscopic blood staining was noted on the tissue contact portion of the device as expected (the extent of which varied depending on the amount of residual blood present at the tissue surface), tissue debris or tissue fragments were not visually observed on the pen tip nor within the tubing system by the surgical or research team.

**Figure S6.**
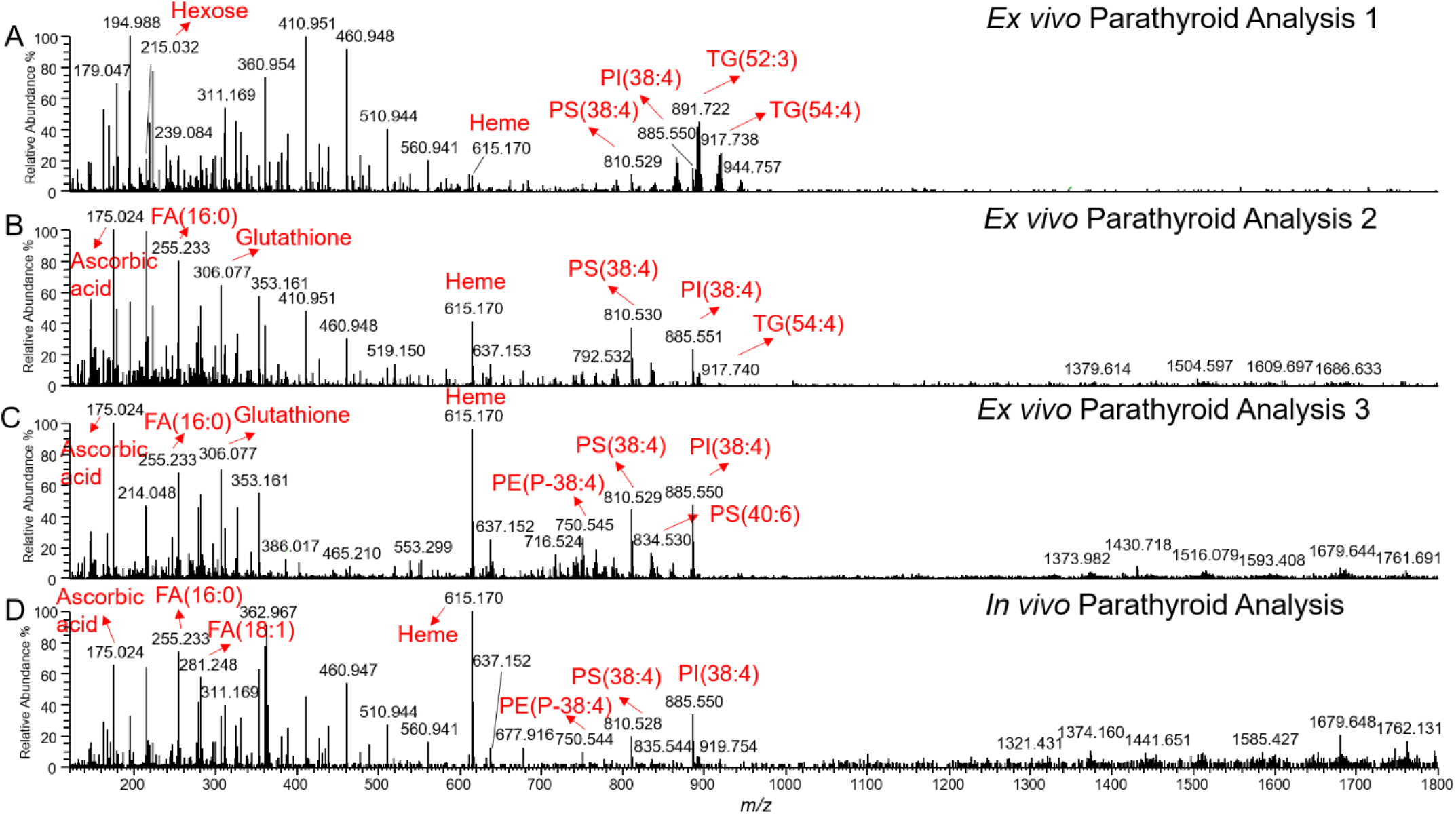
Comparison between mass spectra obtained *ex vivo* (A-C) and *in vivo* (D) from a parathyroid collected with the MasSpec Pen during a parathyroidectomy procedure for primary hyperparathyroidism (PT0010). Higher relative abundances of triacylglyceride species was observed in the first analysis, while a higher relative abundances of various metabolites, including ascorbic acid and glutathione, as well as glycerophosphoethanolamine, glycerophosphoserine, and glycophosphoinositol lipid species, among others, were detected in the second and third analyses.

**Figure S7.**
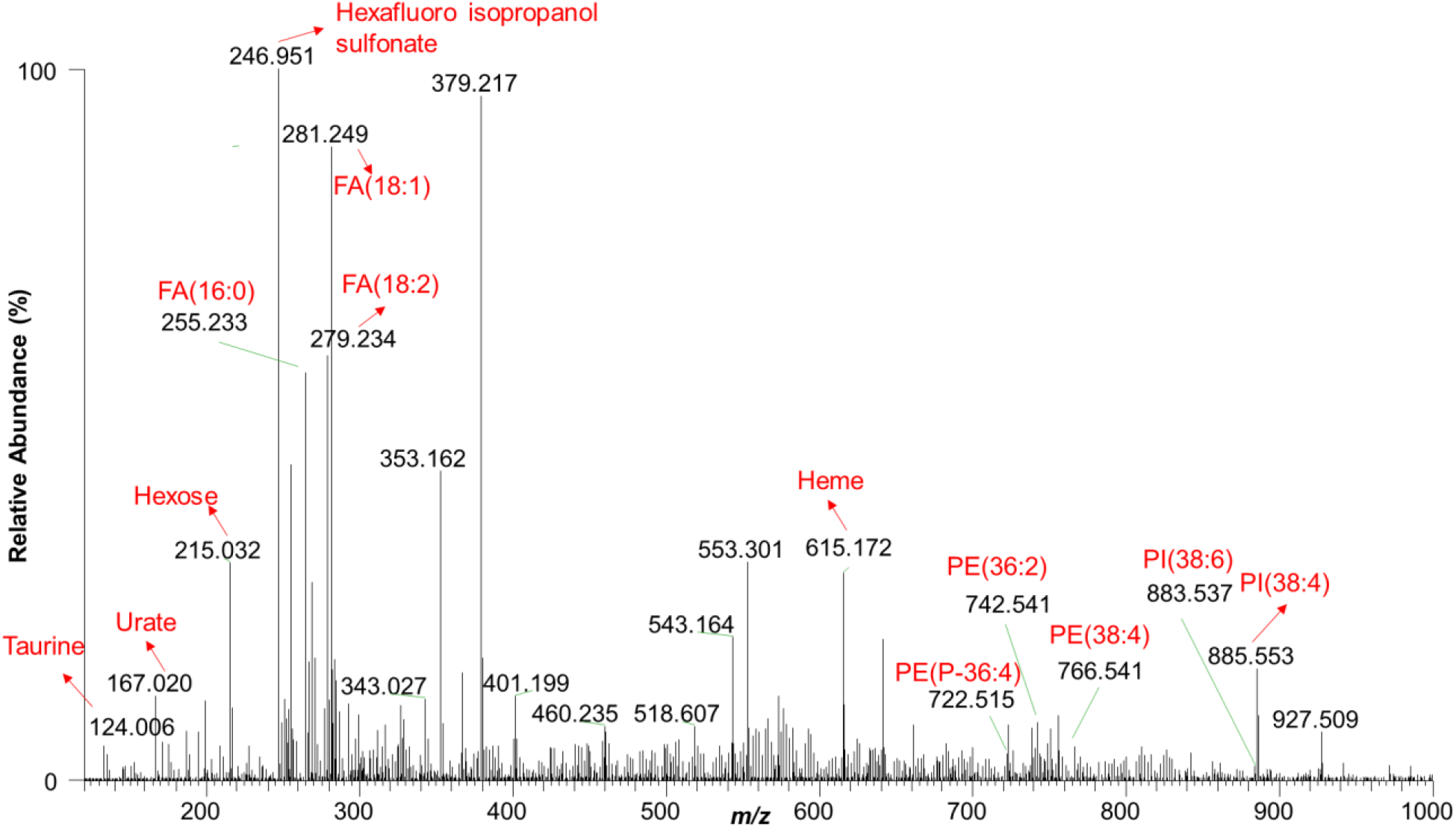
*Ex vivo* mass spectrum obtained from MasSpec Pen analysis of an excised lymph node sample during a right mastectomy procedure with sentinel node biopsy on a patient with grade III intraductal carcinoma (BR0001).

**Figure S8.**
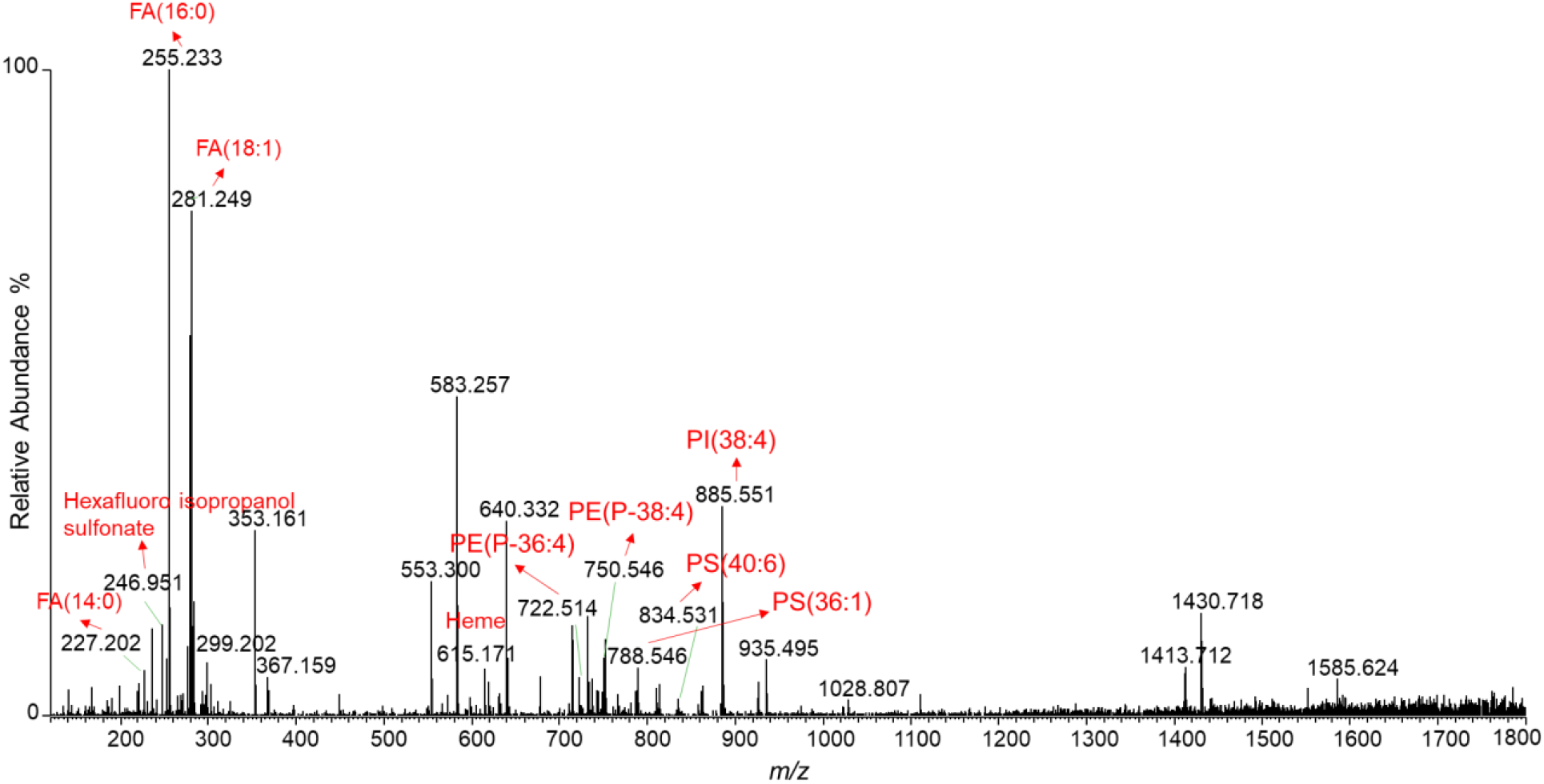
Mass spectrum obtained from MasSpec Pen *ex vivo* analysis of an excised breast specimen during a second surgery right breast lumpectomy for re-excision of a positive ductal carcinoma in situ margin (BR0003).

**Figure S9.**
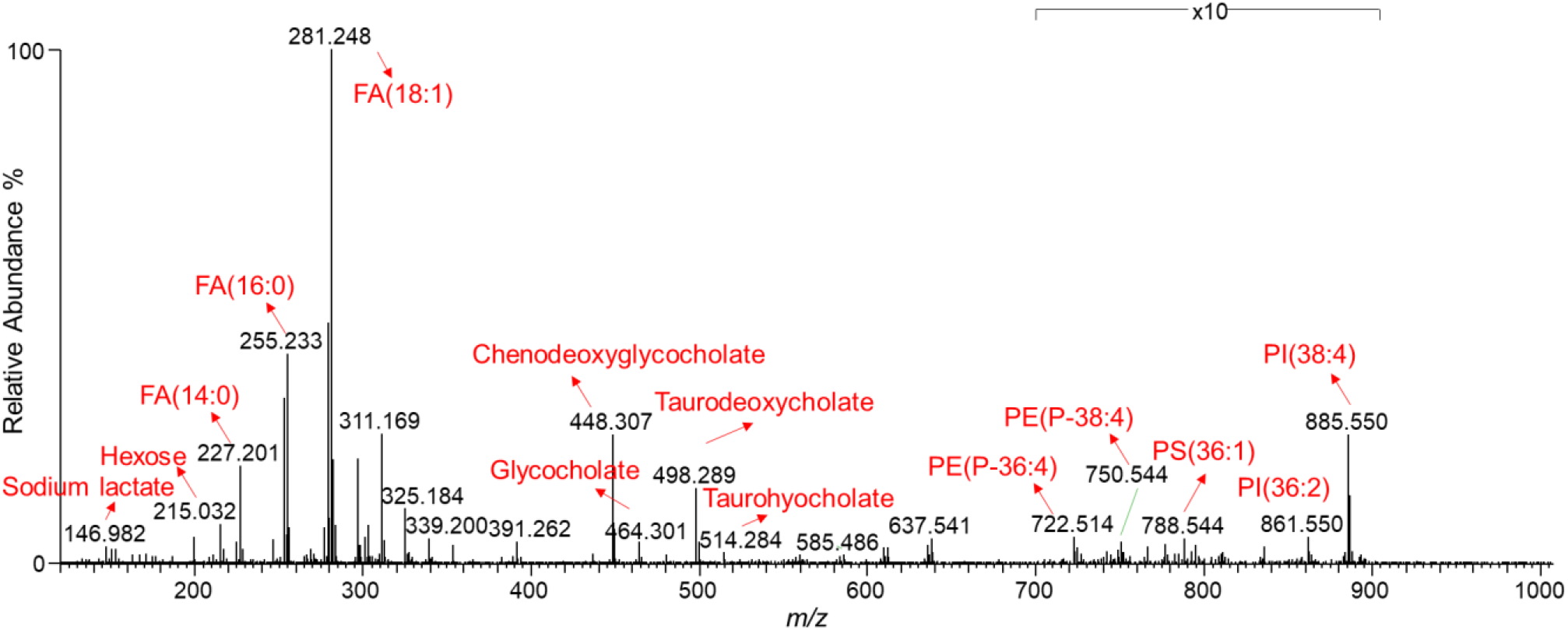
*Ex vivo* mass spectrum of an excised pancreatic specimen collected with the MasSpec Pen during a Whipple procedure for a pancreatic mass (PN0006).

**Figure S10.**
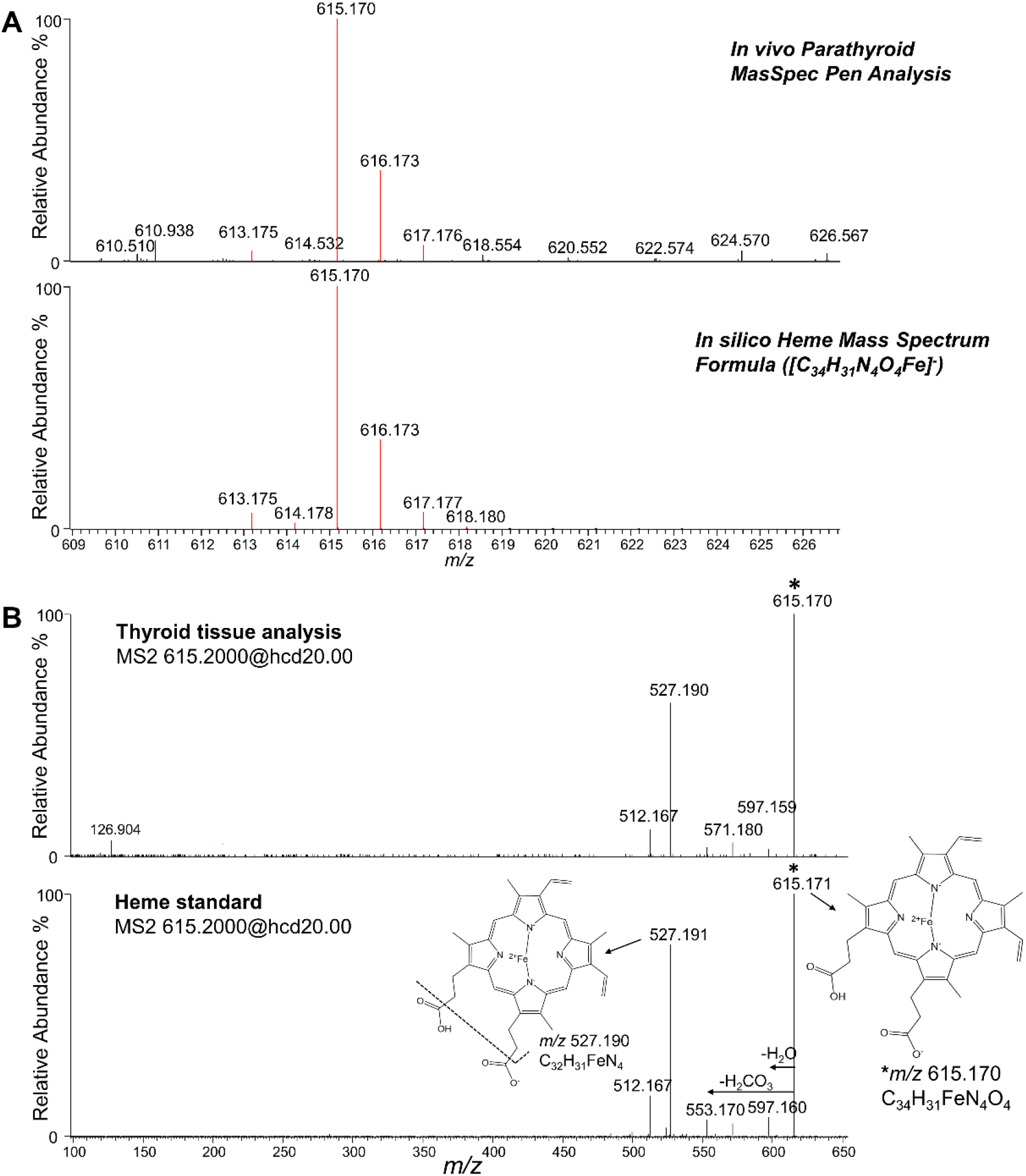
(A) Isotopic distribution of heme (on the top) observed *in vivo* from MasSpec Pen analysis of parathyroid tissue during a parathyroidectomy procedure for primary hyperparathyroidism (PT0012) and theoretical mass spectrum of heme (at the bottom) in negative ion mode using isotope simulation function available in Thermo XCalibur Qual Browser. (B) Tandem mass spectrum of *m/z* 615.170 (on the top) from a thyroid tissue analysis by the MasSpec Pen and tandem mass spectrum of *m/z* 615.171 (at the bottom) from heme standard by electrospray ionization (ESI) under higher-energy collisional dissociation (HCD) mode.

**Figure S11.**
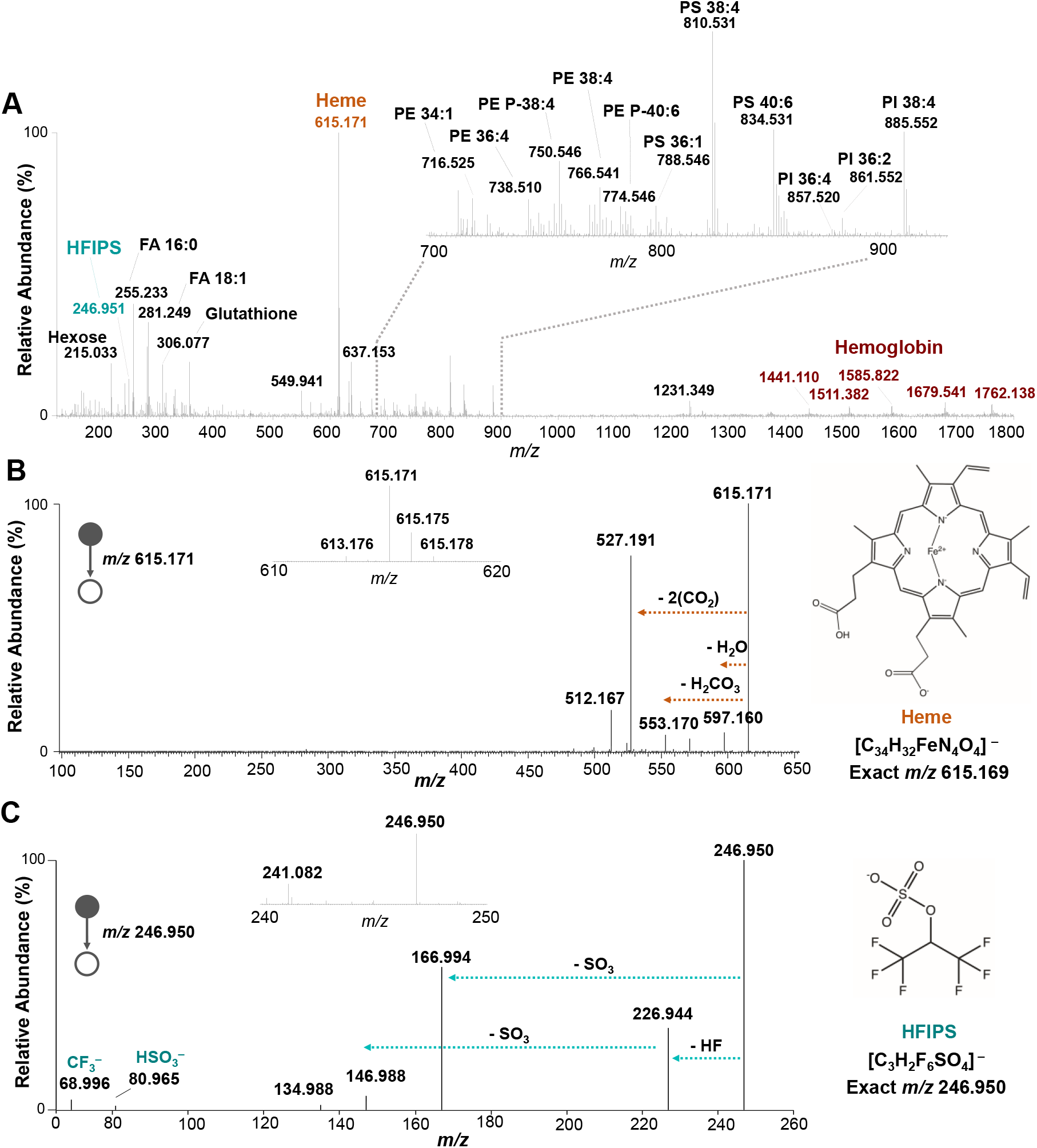
Detection and identification of heme and hexafluoroisopropyl sulfonate. (**A**) *In vivo* mass spectrum obtained from MasSpec Pen analysis of a normal parathyroid specimen during parathyroidectomy procedure for primary hyperparathyroidism (PT0020). Despite the detection of heme at the highest relative abundance (100%) and the detection of multiply charged hemoglobin peaks, various metabolite and lipid species were still detected and identified, as annotated for selected ions in the mass spectra. (**B**) Identification of *m/z* 615.171 as deprotonated heme by MS^2^ experiment performed during analysis of a thyroid tissue by the MasSpec Pen. (**C**) Identification of *m/z* 246.950 as deprotonated hexafluoroisopropyl sulfonate (HFIPS) by MS^2^ performed on tissue.

**Figure S12.**
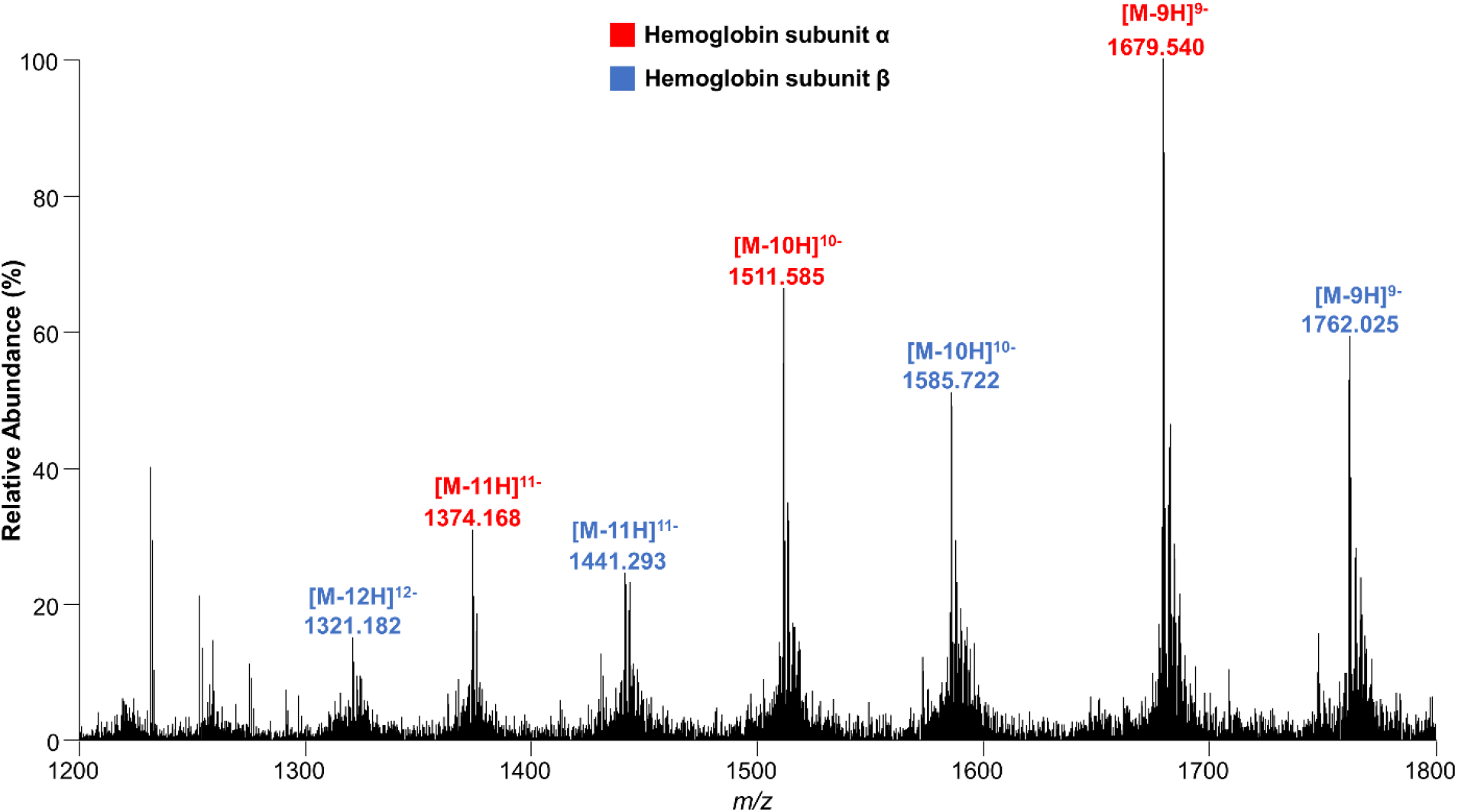
Multiply charged peaks detected from *in vivo* analysis of thyroid tissue using MasSpec Pen during a right hemithyroidectomy procedure of an indeterminate nodule (TH0025) were tentatively assigned as hemoglobin α and hemoglobin β subunits. Note that monoisotopic masses of human hemoglobin α and hemoglobin β subunits (without the initiator methionine) are 15116 and 15857, respectively.

**Figure S13.**
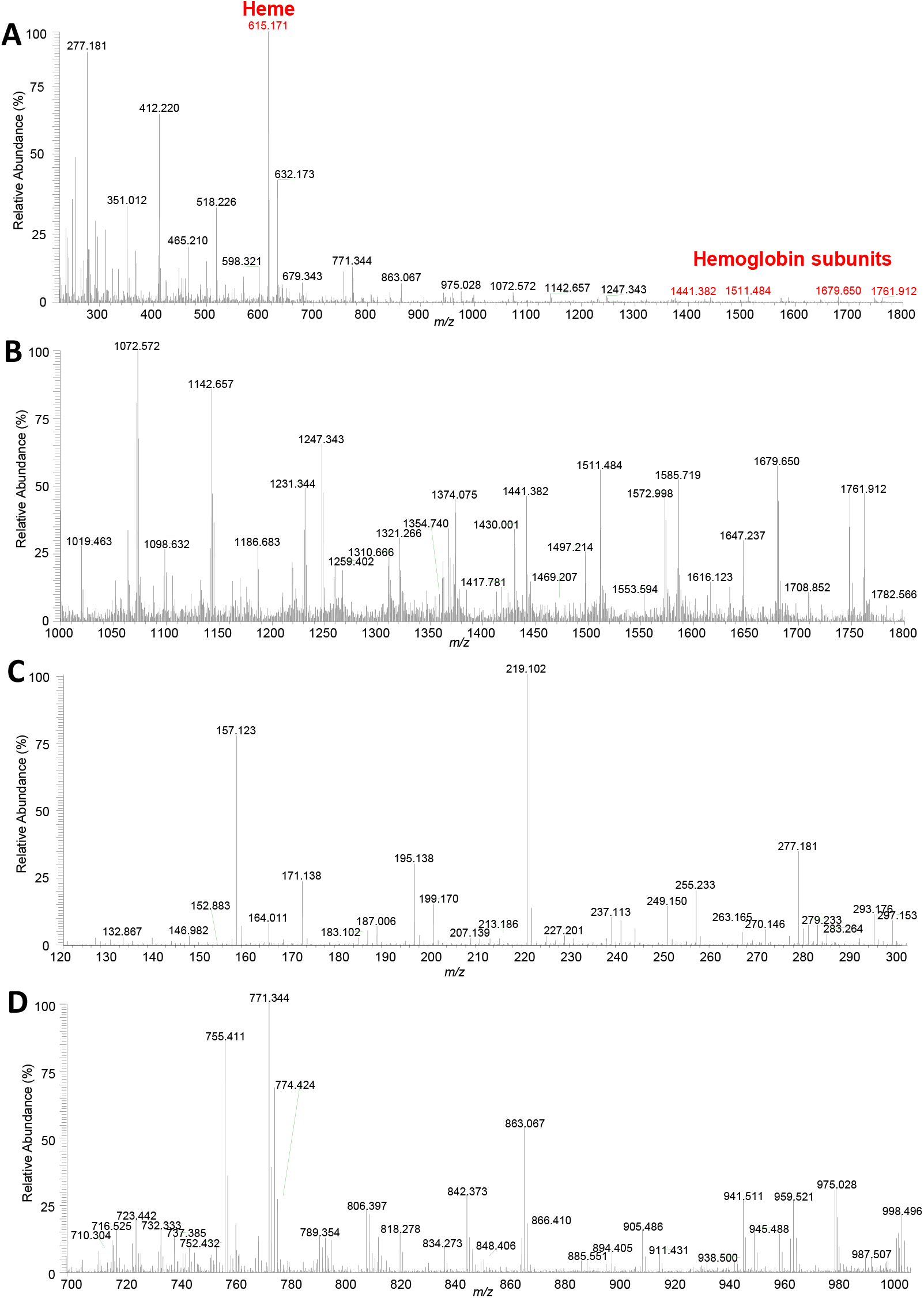
Negative ion mode mass spectra obtained of 10 µL of human blood deposited onto a glass slide and analyzed with the MasSpec Pen. (A) *m/z* 220-1800 range showing detection of heme and hemoglobin peaks, (B) zoom in *m/z* 1000-1800 showing detecting of multiply charged hemoglobin peaks, (C) zoom in *m/z* 120-300 where metabolites and fatty acids are normally detected, (D) zoom in *m/z* 700-1000 where complex lipids are often detected.

**Figure S14.**
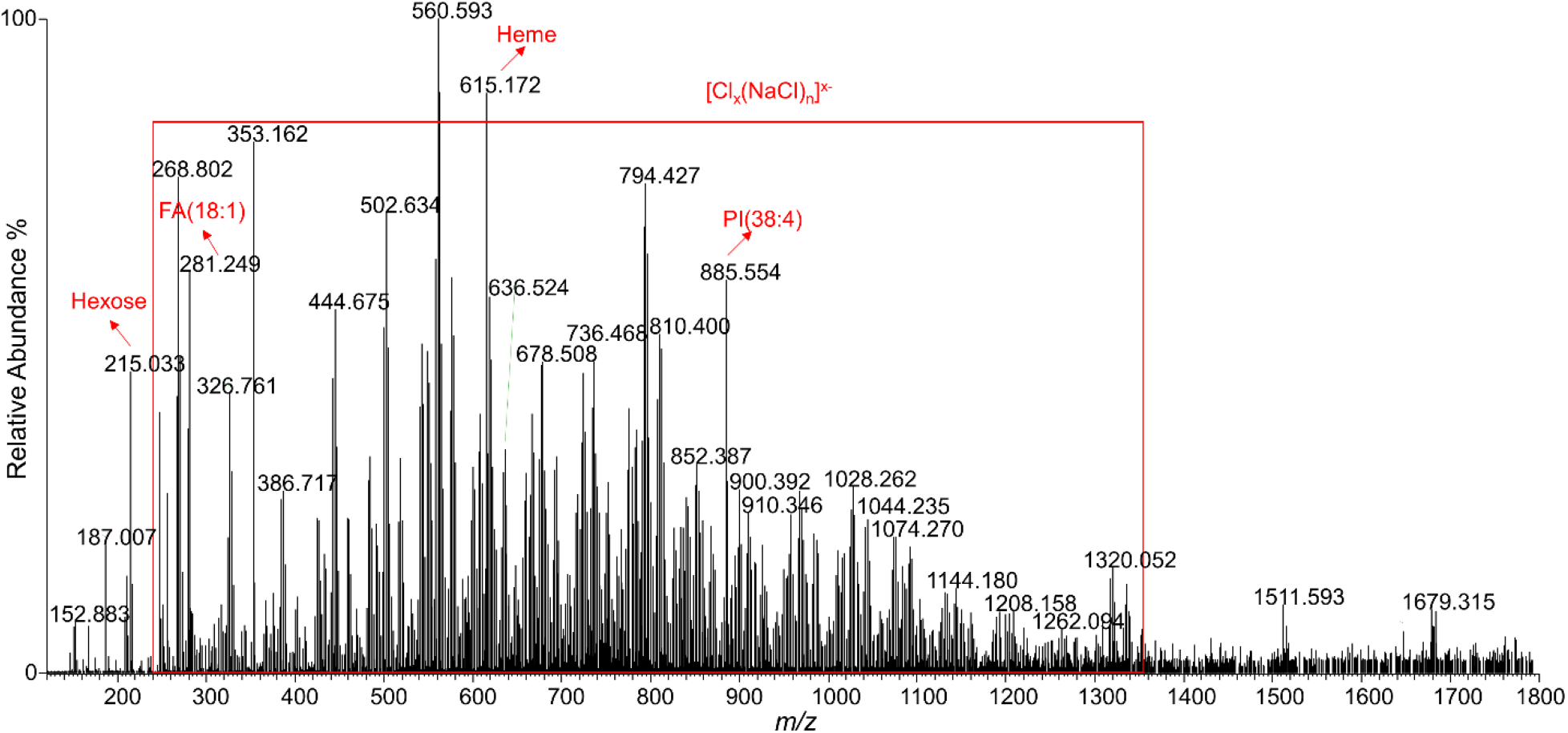
MasSpec Pen *in vivo* analysis of a thyroid nodule during a total thyroidectomy and parathyroidectomy for a thyroid neoplasm and primary hyperparathyroidism (TH0024), showing interferences in the mass spectrum resulting from saline irrigation

**Figure S15.**
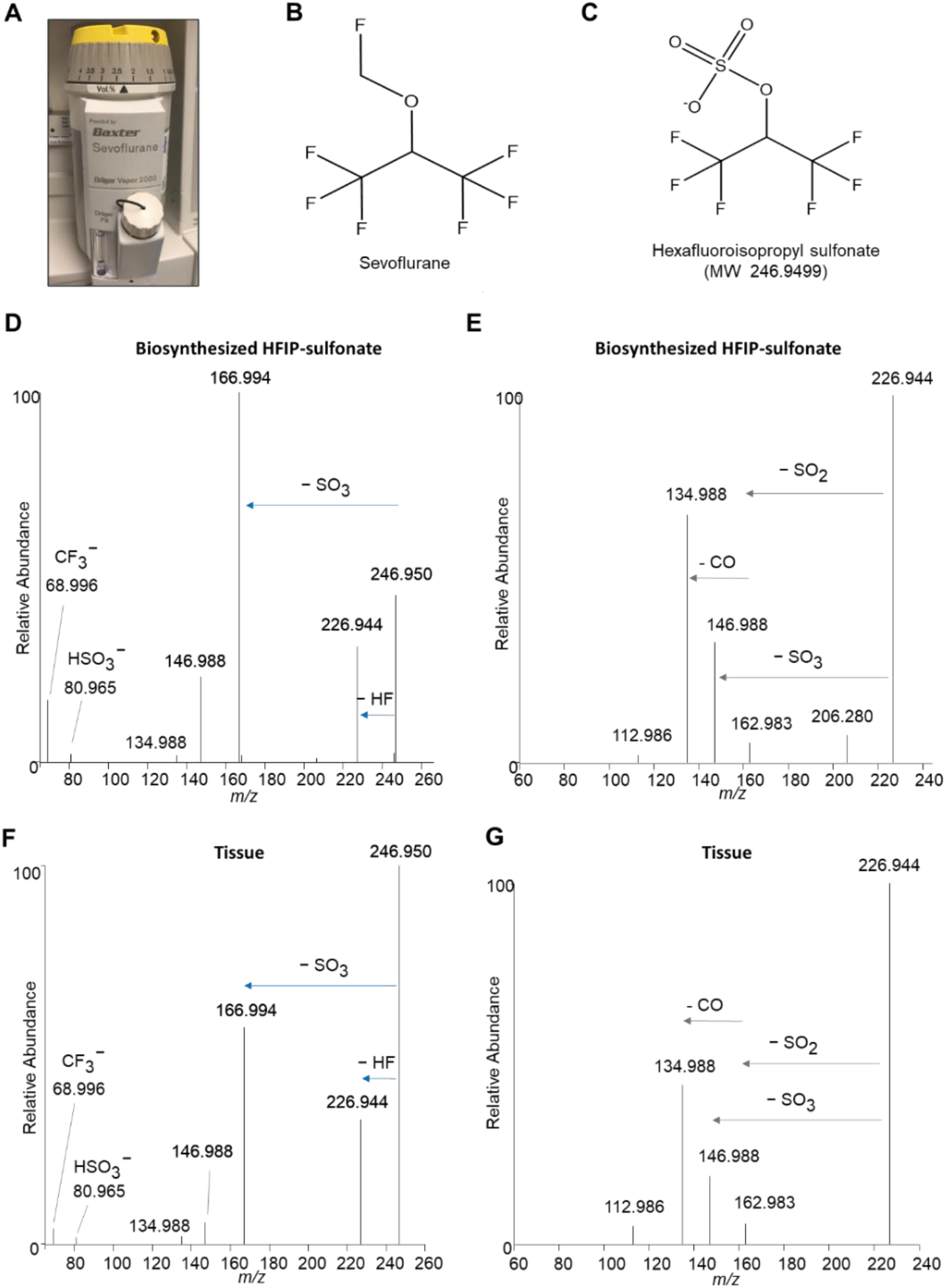
Identification of *m/z* 246.950 as hexafluoroisopropyl sulfonate (HFIPS). (A) Sevofluorane tank used for anesthesia during the surgeries described in this study. (B) Structure of sevofluorane. (C) Structure of HFIPS. MS^2^ (D) and MS^3^ (E) of biosynthesized HFIPS. MS^2^ (F) and MS^3^ (G) of HFIPS detected from endometriosis tissue by DESI-MS.

## Supplementary Tables

**Table S1.** Cosine similarity results between the mass spectra obtained with different tubing lengths of the MasSpec Pen.

| Tubing Length |  |  |
| --- | --- | --- |
|  | 1.5 meters (n=4) | 3.0 meters (n=4) |
| 3.0 meters (n=4) | 0.91 | -- |
| 4.5 meters (n=4) | 0.92 | 0.97 |

**Table S2.** Droplet transport time from pen tip to mass spectrometer through various tubing lengths.

| Droplet Transport Time |  |  |
| --- | --- | --- |
|  | Average (n=10)<br>in seconds | Relative Standard Deviation (RSD) |
| 1.5 meters | 3.8 | 12.4% |
| 3.0 meters | 5.8 | 11.5% |
| 4.5 meters | 7.5 | 5.2% |

**Table S3.**
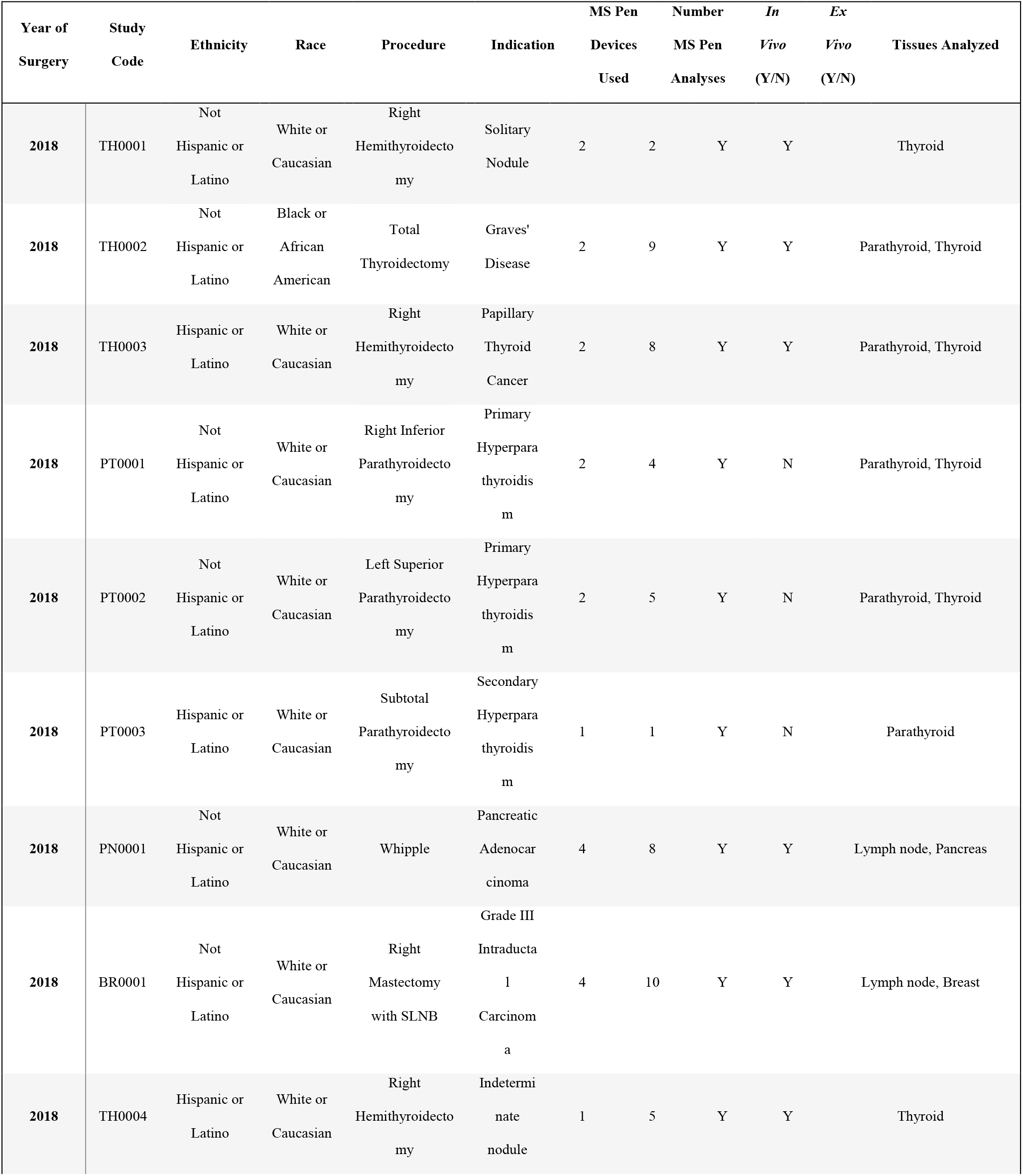
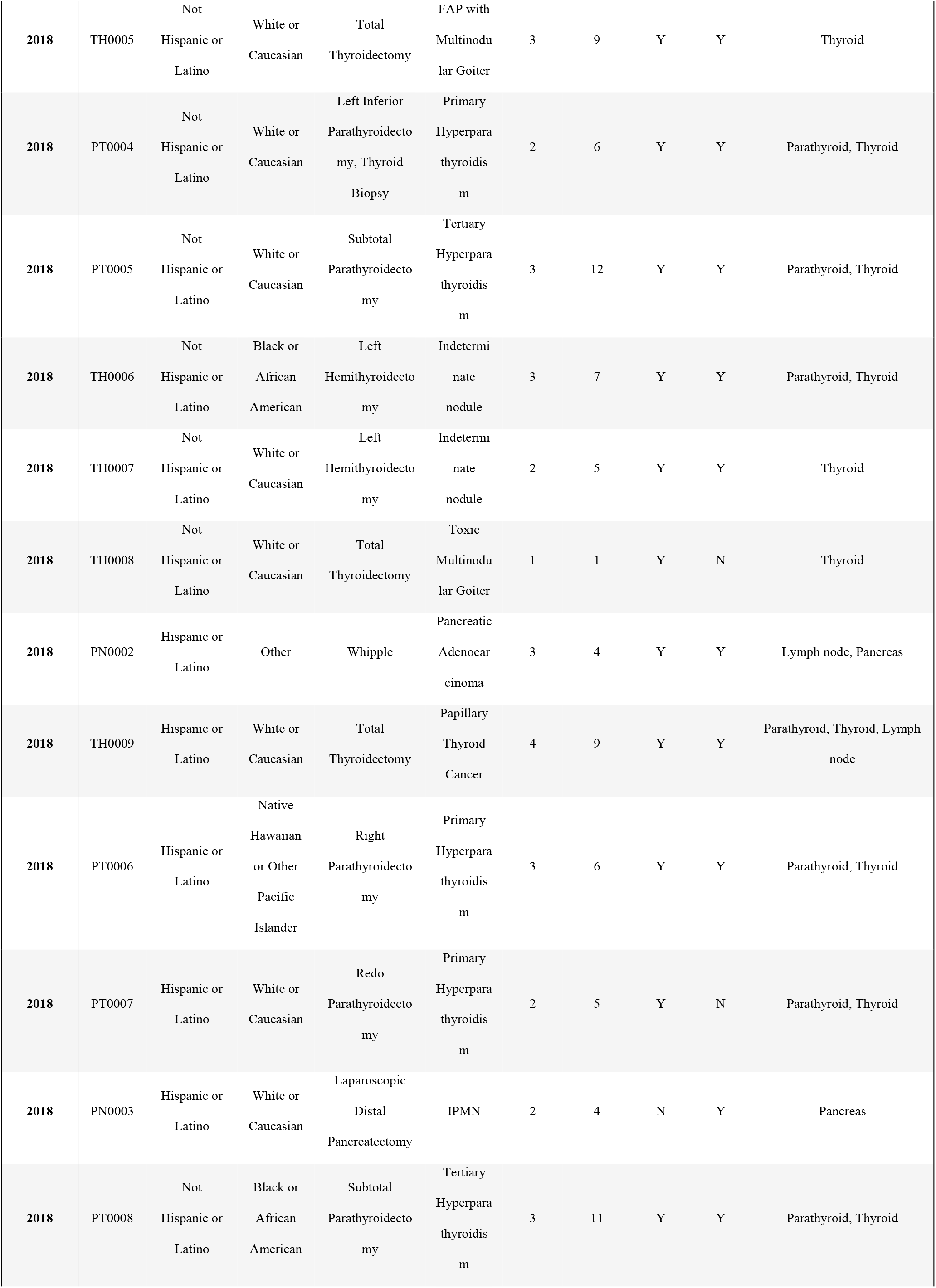
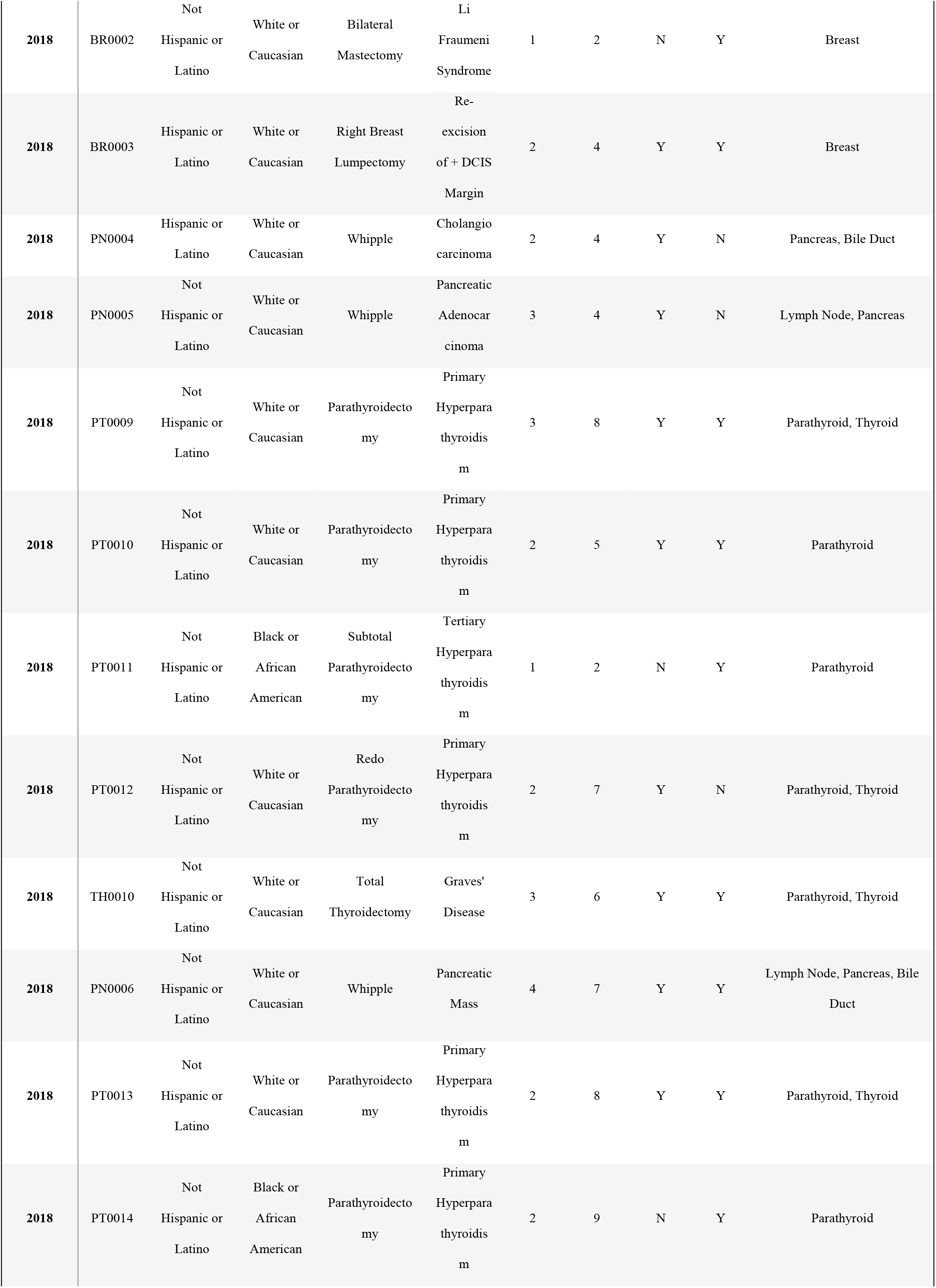
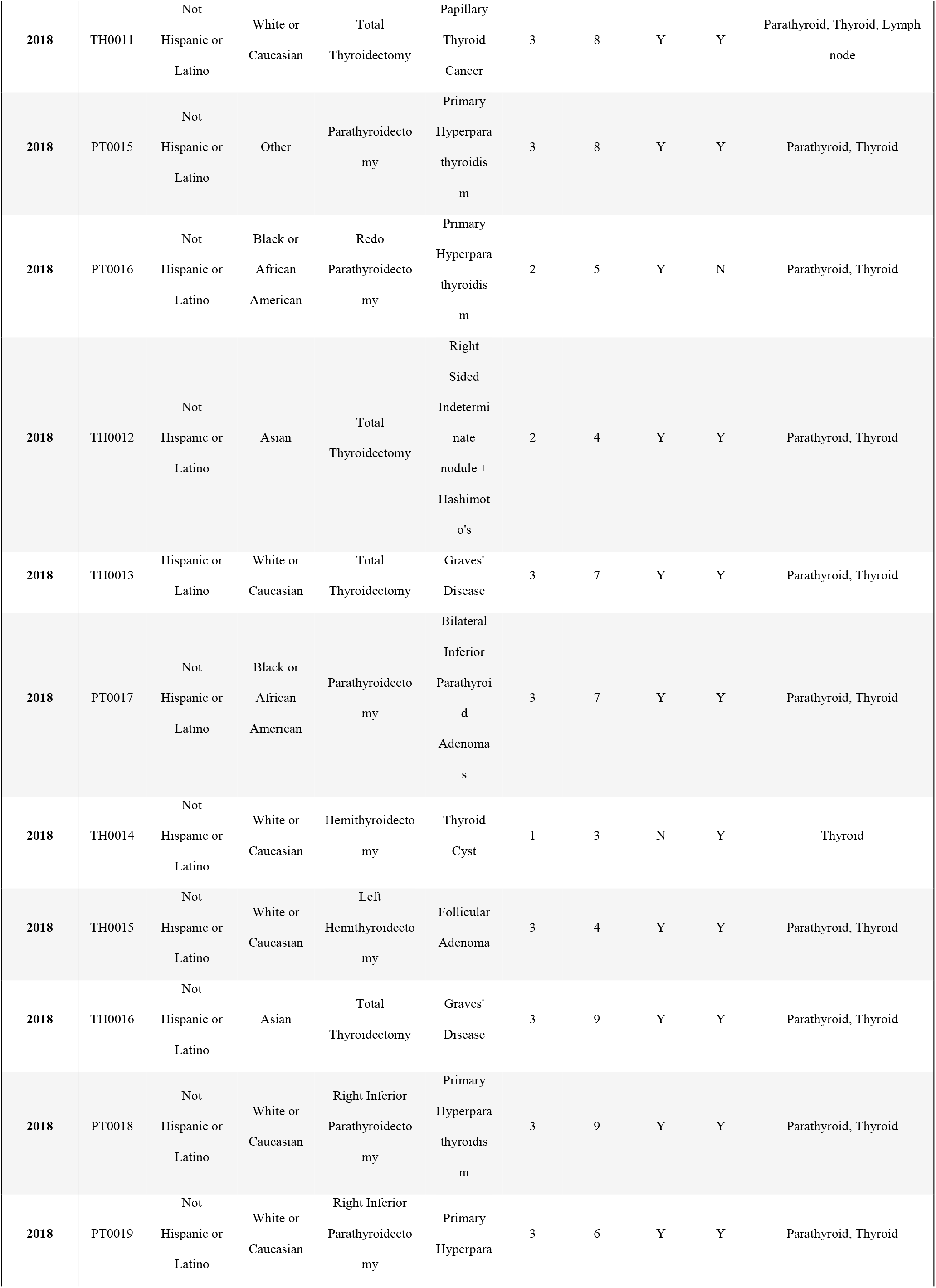
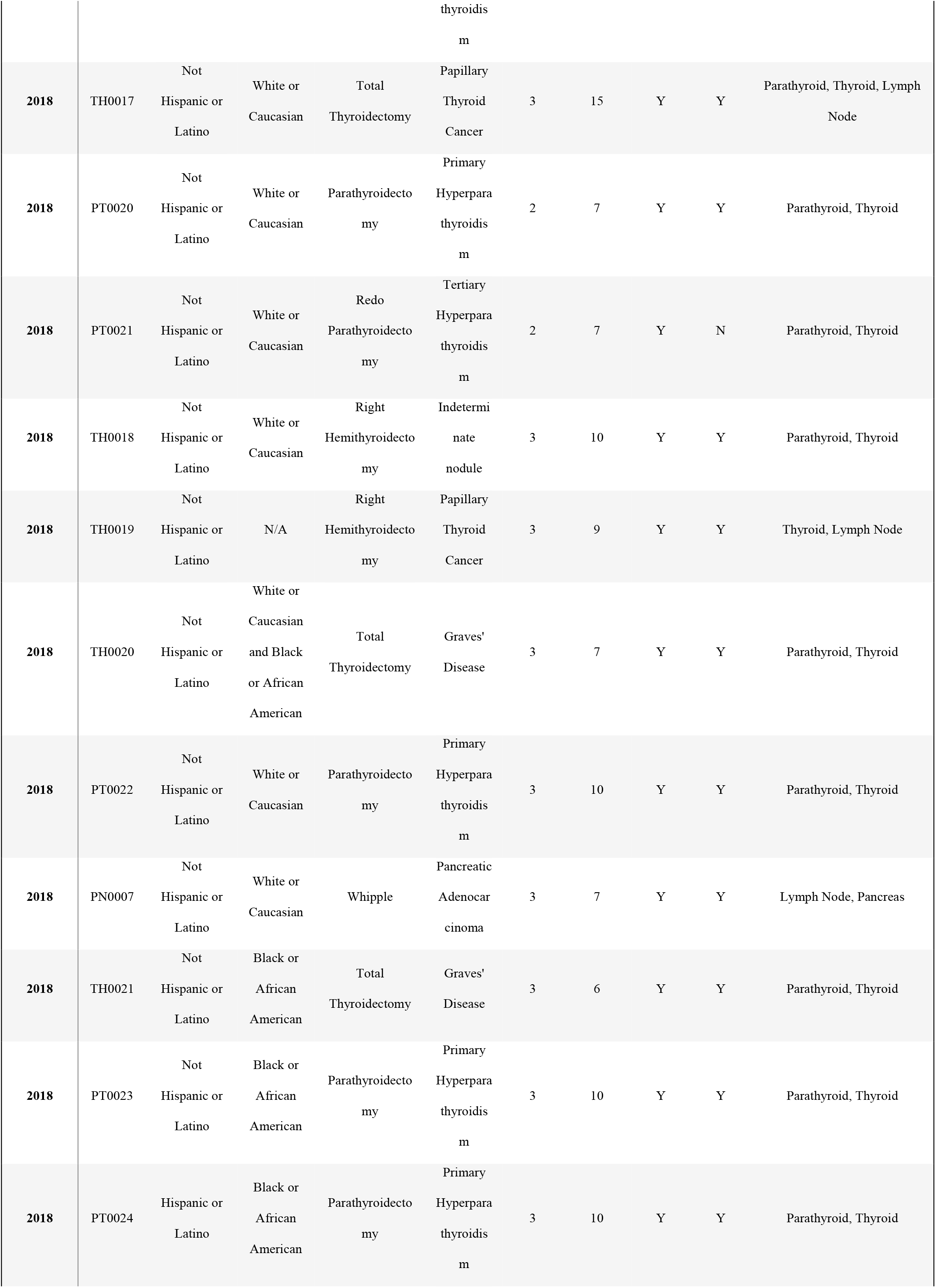
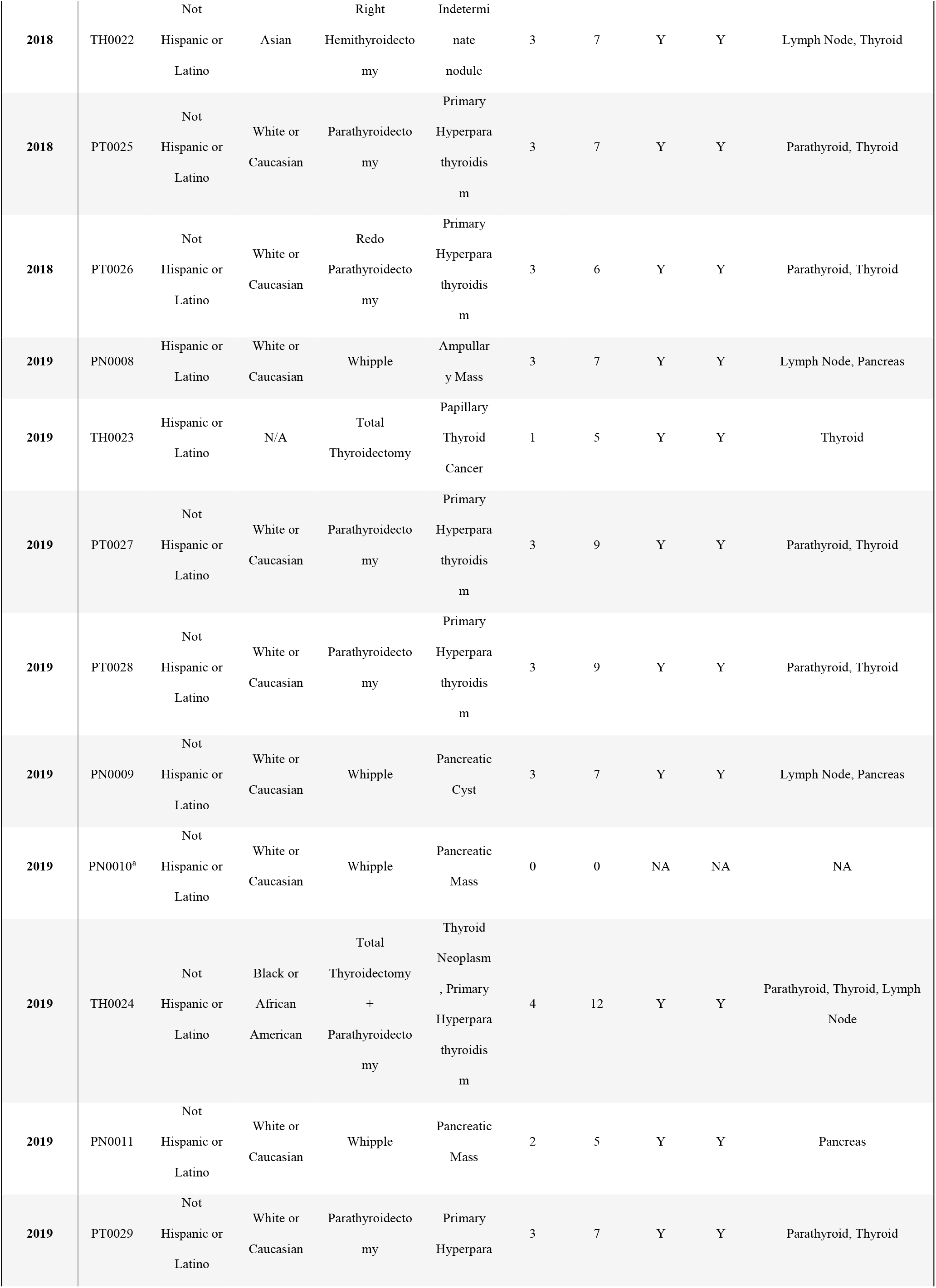
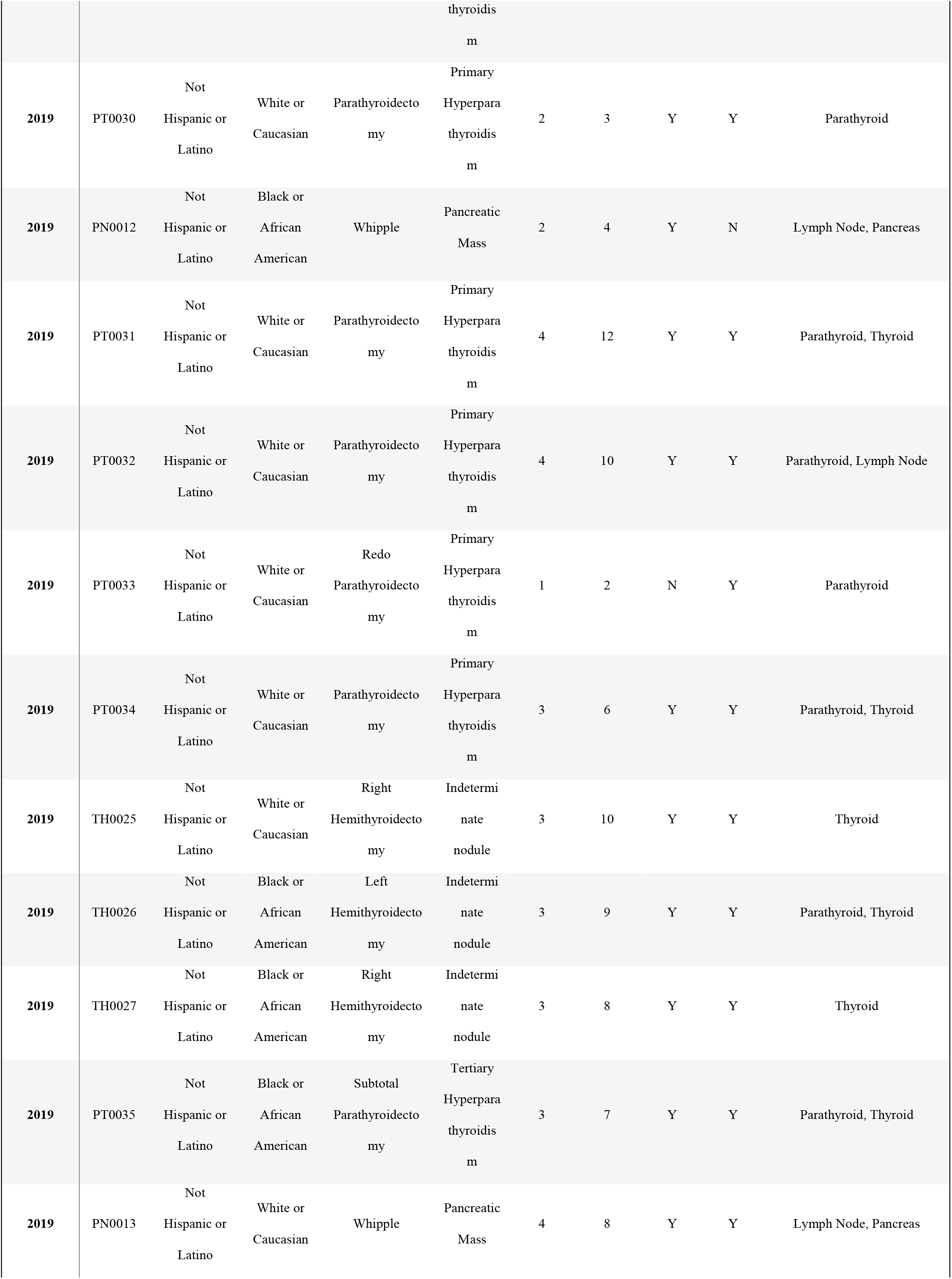
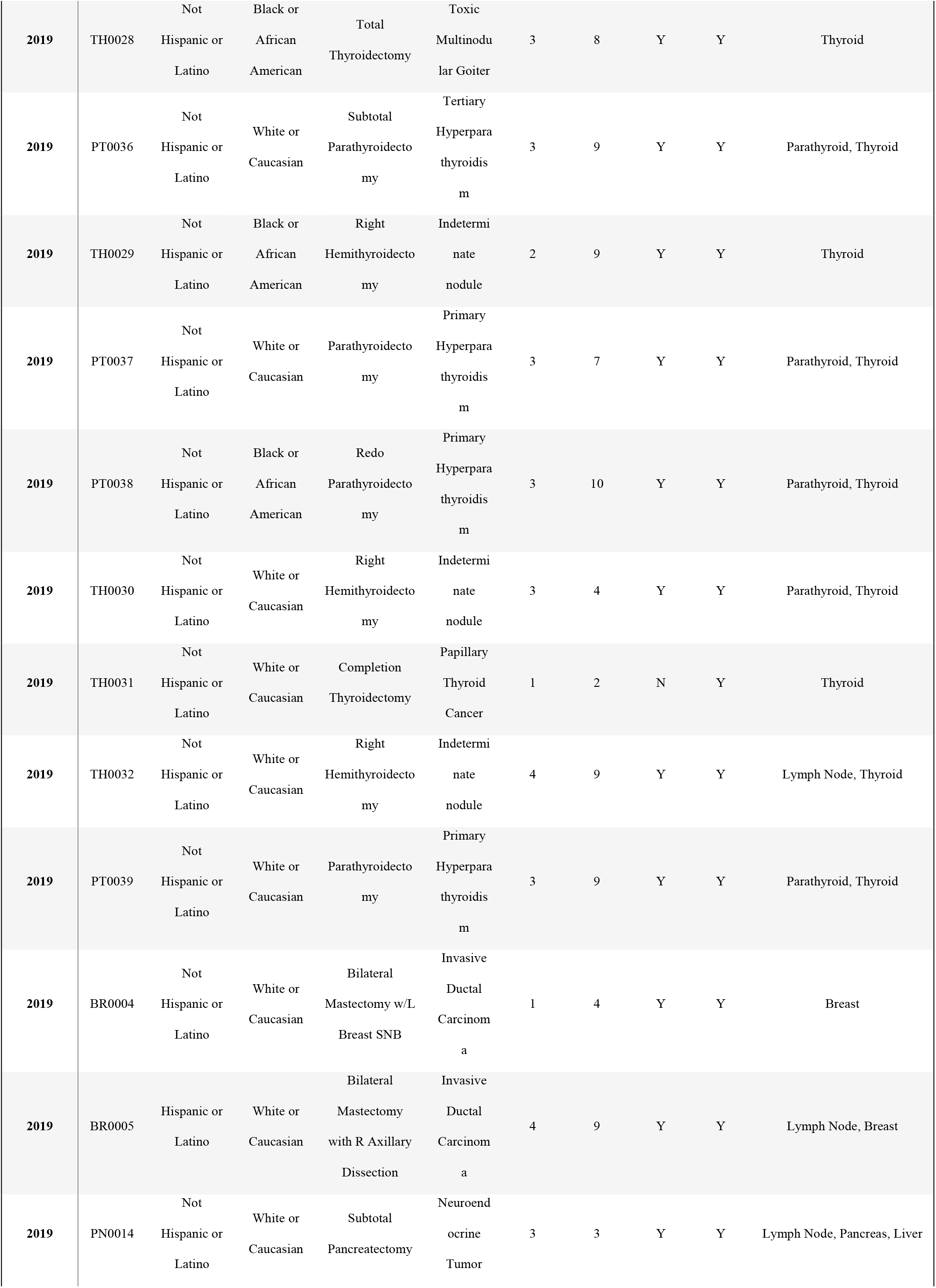
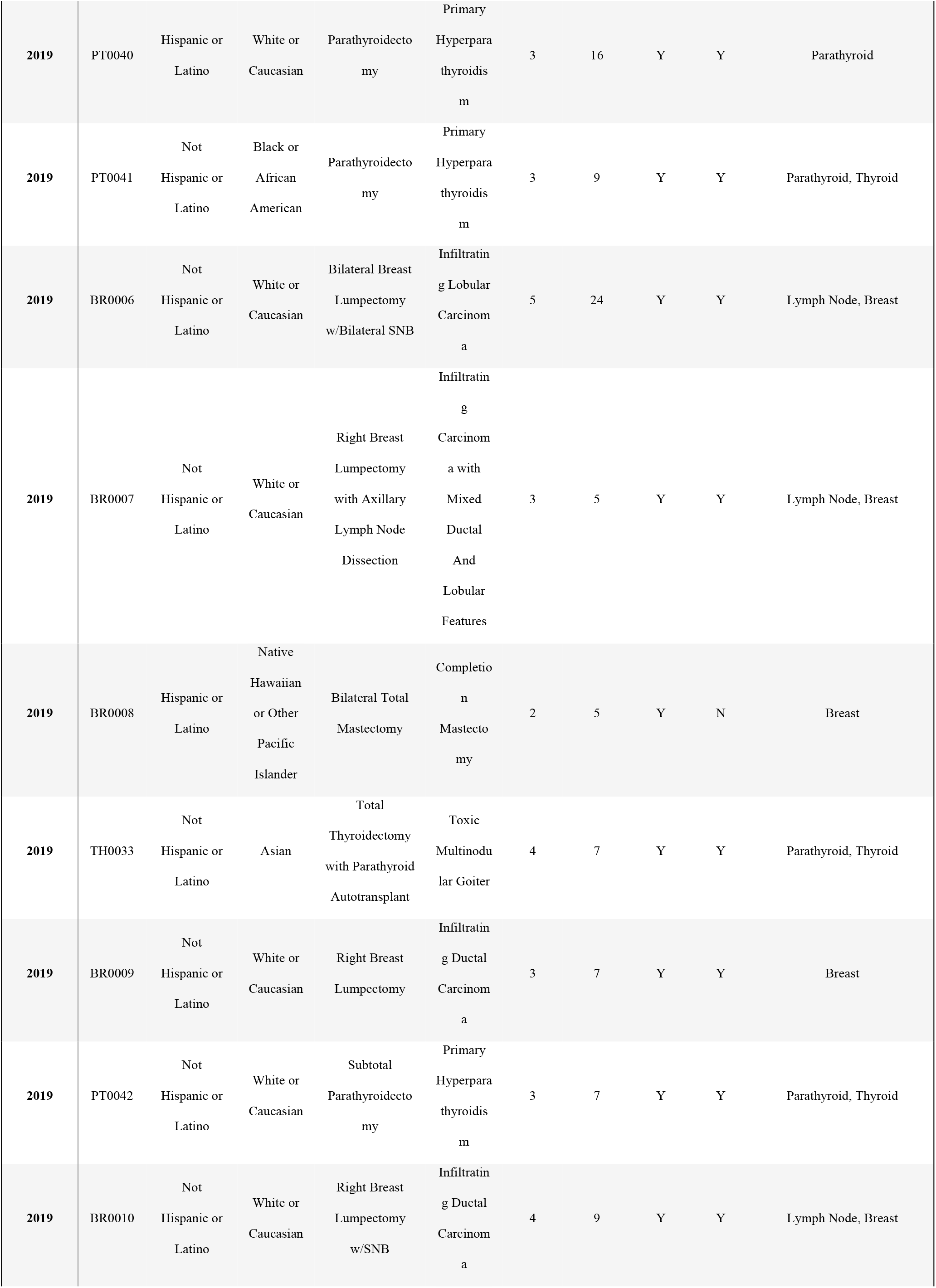
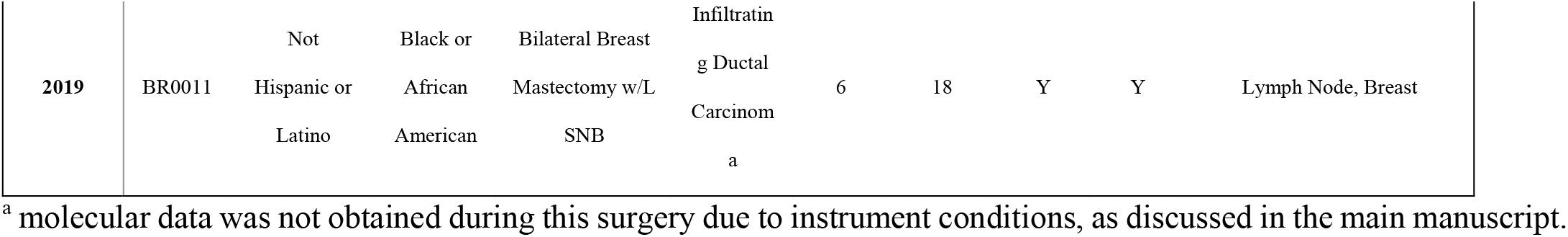
Clinical and demographic information for the 100 patients included in this study. Information on the analyses conducted with the MasSpec Pen, such as number of devices used, number of analyses, tissue samples analyzed and type of analysis (*in vivo* or *ex vivo*), is also provided. Patients IDs were defined according to the type of procedure (thyroidectomies – TH, parathyroidectomies – PT, BR – breast cancer surgery, PN – pancreatic cancer surgery).

**Table S4.** Proposed attributions, molecular formula, and mass error for the species annotated. Abbreviations: FA – fatty acid, PE – glycerophosphoethanolamine, PS – glycerophosphoserine, PI – glycerophosphoinositol, TG – triacylglycerol.

| Detected $m/z$ | Proposed Attribution | Proposed Molecular Formula | Mass Error (ppm) |
| --- | --- | --- | --- |
| 124.006 | Taurine | $C_2H_6NO_3S$ | 3.6 |
| 126.904 | Iodine | I | 7.1 |
| 146.045 | Glutamic Acid | $C_5H_8NO_4$ | 5.5 |
| 167.021 | Uric Acid | $C_5H_3N_4O_3$ | 3.0 |
| 175.024 | Ascorbic Acid | $C_6H_7O_6$ | 2.3 |
| 191.020 | Citric Acid | $C_6H_7O_7$ | 1.0 |
| 215.033 | Hexose | $C_6H_{12}O_6Cl$ | -0.5 |
| 246.951 | Hexafluoroisopropyl sulfonate | $C_3HO_4F_6S$ | -2.4 |
| 227.202 | FA 14:0 | $C_{14}H_{27}O_2$ | 1.5 |
| 255.234 | FA 16:0 | $C_{16}H_{31}O_2$ | -2.7 |
| 279.234 | FA 18:2 | $C_{18}H_{31}O_2$ | -3.6 |
| 281.250 | FA 18:1 | $C_{18}H_{33}O_2$ | -3.9 |
| 283.265 | FA 18:0 | $C_{18}H_{35}O_2$ | -3.9 |
| 303.234 | FA 20:4 | $C_{20}H_{31}O_2$ | -4.3 |
| 306.078 | Glutathione | $C_{10}H_{16}N_3O_6S$ | -4.6 |
| 448.307 | Chenodeoxyglycocholate | $C_{26}H_{42}O_5N$ | 0.3 |
| 464.302 | Glycocholate | $C_{26}H_{42}O_6N$ | 0.5 |
| 498.290 | Taurodeoxycholate | $C_{26}H_{44}O_6NS$ | 1.0 |
| 514.285 | Taurohyocholate | $C_{26}H_{44}O_7NS$ | 1.2 |
| 543.164 | Lymphazurin | $C_{27}H_{31}N_2O_6S_2$ | 1.8 |
| 615.173 | Heme | $C_{34}H_{31}FeN_4O_4$ | -5.0 |
| 698.514 | PE P-34:2 | $C_{39}H_{73}NO_7P$ | -2.0 |
| 700.532 | PE P-34:1 | $C_{39}H_{75}NO_7P$ | 4.8 |
| 714.511 | PE 34:2 | $C_{39}H_{73}NO_8P$ | 4.3 |
| 716.527 | PE 34:1 | $C_{39}H_{75}NO_8P$ | 4.8 |
| 722.517 | PE P-36:4 | C<sub>41</sub>H<sub>73</sub>NO<sub>7</sub>P | -5.3 |
| 726.549 | PE P-36:2 | C<sub>41</sub>H<sub>77</sub>NO<sub>7</sub>P | -5.8 |
| 738.512 | PE 36:4 | C<sub>41</sub>H<sub>73</sub>NO<sub>8</sub>P | 5.5 |
| 742.543 | PE 36:2 | C<sub>41</sub>H<sub>77</sub>NO<sub>8</sub>P | -5.5 |
| 744.560 | PE 36:1 | C<sub>41</sub>H<sub>79</sub>NO<sub>8</sub>P | 6.9 |
| 750.549 | PE P-38:4 | C<sub>43</sub>H<sub>77</sub>NO<sub>7</sub>P | -6.0 |
| 764.528 | PE 38:5 | C<sub>43</sub>H<sub>75</sub>NO<sub>8</sub>P | 5.8 |
| 766.543 | PE 38:4 | C<sub>43</sub>H<sub>77</sub>NO<sub>8</sub>P | -5.3 |
| 770.575 | PE 38:2 | C<sub>43</sub>H<sub>81</sub>NO<sub>8</sub>P | 5.8 |
| 778.580 | PE P-40:4 | C<sub>45</sub>H<sub>81</sub>NO<sub>7</sub>P | 5.6 |
| 786.533 | PS 36:2 | C<sub>42</sub>H<sub>77</sub>NO<sub>10</sub>P | -5.2 |
| 788.549 | PS 36:1 | C<sub>42</sub>H<sub>79</sub>NO<sub>10</sub>P | -5.1 |
| 794.575 | PE 40:4 | C<sub>45</sub>H<sub>81</sub>NO<sub>8</sub>P | 5.6 |
| 810.533 | PS 38:4 | C<sub>44</sub>H<sub>77</sub>NO<sub>10</sub>P | -5.2 |
| 834.533 | PS 40:6 | C<sub>46</sub>H<sub>77</sub>NO<sub>10</sub>P | -4.8 |
| 835.537 | PI 34:1 | C<sub>43</sub>H<sub>80</sub>O<sub>13</sub>P | -3.5 |
| 836.548 | PS 40:5 | C<sub>46</sub>H<sub>79</sub>NO<sub>10</sub>P | 3.9 |
| 857.523 | PI 36:4 | C<sub>45</sub>H<sub>78</sub>O<sub>13</sub>P | -4.8 |
| 861.554 | PI 36:2 | C<sub>45</sub>H<sub>82</sub>O<sub>13</sub>P | -5.1 |
| 883.539 | PI 38:5 | C<sub>47</sub>H<sub>80</sub>O<sub>13</sub>P | 5.4 |
| 885.555 | PI 38:4 | C<sub>47</sub>H<sub>82</sub>O<sub>13</sub>P | -5.5 |
| 891.723 | TG 52:3 | C<sub>55</sub>H<sub>100</sub>O<sub>6</sub>Cl | 1.8 |
| 917.739 | TG 54:4 | C<sub>57</sub>H<sub>102</sub>O<sub>6</sub>Cl | 2.1 |

## Notes

### Author Declarations

The study was reviewed and approved by the Institutional Review Board (IRB) of Baylor College of Medicine and the IRB of the University of Texas at Austin

## References

1. Y. Yokoyama, Y. Nimura, M. Nagino, Advances in the treatment of pancreatic cancer: Limitations of surgery and evaluation of new therapeutic strategies. Surg. Today 39, 466–475 (2009).

2. H. Jaafar, Intra-Operative Frozen Section Consultation: Concepts, Applications and Limitations. Malays. J. Med. Sci. 13, 4–12 (2006).

3. K. Lee et al., The potential role of frozen sections of tumors in decision making of axillary procedure in breast conserving surgery for DCIS at preoperative diagnosis. Breast 44, S119–S119 (2019).

4. R. K. Orosco et al., Positive Surgical Margins in the 10 Most Common Solid Cancers. Sci. Rep. 8 (2018).

5. A. A. Gal, P. T. Cagle, The 100-year anniversary of the description of the frozen section procedure. J. Am. Med. Assoc. 294, 3135–3137 (2005).

6. T. A. Buchholz et al., Margins for Breast-Conserving Surgery With Whole-Breast Irradiation in Stage I and II Invasive Breast Cancer: American Society of Clinical Oncology Endorsement of the Society of Surgical Oncology/American Society for Radiation Oncology Consensus Guideline. J. Clin. Oncol. 32, 1502–1506 (2014).

7. M. S. Moran et al., Society of Surgical Oncology-American Society for Radiation Oncology Consensus Guideline on Margins for Breast-Conserving Surgery With Whole-Breast Irradiation in Stages I and II Invasive Breast Cancer. J. Clin. Oncol. 32, 1507–1515 (2014).

8. T. A. King et al., Clinical Management Factors Contribute to the Decision for Contralateral Prophylactic Mastectomy. J. Clin. Oncol. 29, 2158–2164 (2011).

9. D. T. Hughes et al., Influence of prophylactic central lymph node dissection on postoperative thyroglobulin levels and radioiodine treatment in papillary thyroid cancer. Surgery 148, 1100-1106; discussion 1006-1107 (2010).

10. M. A. Adam et al., Is There a Minimum Number of Thyroidectomies a Surgeon Should Perform to Optimize Patient Outcomes? Ann. Surg. 265, 402–407 (2017).

11. B. S. Koo et al., Predictive factors for ipsilateral or contralateral central lymph node metastasis in unilateral papillary thyroid carcinoma. Ann Surg 249, 840–844 (2009).

12. A. M. Laird, P. G. Gauger, B. S. Miller, G. M. Doherty, Evaluation of Postoperative Radioactive Iodine Scans in Patients who Underwent Prophylactic Central Lymph Node Dissection. World J Surg 36, 1268–1273 (2012).

13. C. Kut et al., Detection of human brain cancer infiltration ex vivo and in vivo using quantitative optical coherence tomography. Sci. Transl. Med. 7 (2015).

14. N. Rajaram et al., Design and validation of a clinical instrument for spectral diagnosis of cutaneous malignancy. Appl. Opt. 49, 142–152 (2010).

15. D. Calligaris et al., MALDI mass spectrometry imaging analysis of pituitary adenomas for near-real-time tumor delineation. Proc. Natl. Acad. Sci. U. S. A. 112, 9978–9983 (2015).

16. S. Banerjee et al., Diagnosis of prostate cancer by desorption electrospray ionization mass spectrometric imaging of small metabolites and lipids. Proc. Natl. Acad. Sci. U. S. A. 114, 3334–3339 (2017).

17. M. Woolman et al., Picosecond Infrared Laser Desorption Mass Spectrometry Identifies Medulloblastoma Subgroups on Intrasurgical Timescales. Cancer Res. 79, 2426–2434 (2019).

18. D. Calligaris et al., Application of desorption electrospray ionization mass spectrometry imaging in breast cancer margin analysis. Proc. Natl. Acad. Sci. U. S. A. 111, 15184–15189 (2014).

19. N. Ogrinc et al., Water-assisted laser desorption/ionization mass spectrometry for minimally invasive in vivo and real-time surface analysis using SpiderMass. Nat. Protoc. 14, 3162–3182 (2019).

20. J. Alexander et al., A novel methodology for in vivo endoscopic phenotyping of colorectal cancer based on real-time analysis of the mucosal lipidome: a prospective observational study of the iKnife. Surg. Endosc. 31, 1361–1370 (2017).

21. M. Jermyn et al., Intraoperative brain cancer detection with Raman spectroscopy in humans. Sci. Transl. Med. 7, 274ra219 (2015).

22. D. A. Orringer et al., Rapid intraoperative histology of unprocessed surgical specimens via fibre-laser-based stimulated Raman scattering microscopy. Nat. Biomed. Eng. 1, 0027 (2017).

23. M. Woolman et al., Rapid determination of medulloblastoma subgroup affiliation with mass spectrometry using a handheld picosecond infrared laser desorption probe. Chem Sci 8, 6508–6519 (2017).

24. B. Fatou et al., In vivo Real-Time Mass Spectrometry for Guided Surgery Application. Sci Rep 6, 25919 (2016).

25. V. Pirro et al., Intraoperative assessment of tumor margins during glioma resection by desorption electrospray ionization-mass spectrometry. Proc Natl Acad Sci U S A 114, 6700–6705 (2017).

26. M. Sans et al., Performance of the MasSpec Pen for Rapid Diagnosis of Ovarian Cancer. Clin. Chem. 65, 674–683 (2019).

27. L. S. Eberlin et al., Pancreatic Cancer Surgical Resection Margins: Molecular Assessment by Mass Spectrometry Imaging. PLoS Med. 13, e1002108 (2016).

28. J. L. Zhang et al., Nondestructive tissue analysis for ex vivo and in vivo cancer diagnosis using a handheld mass spectrometry system. Sci. Transl. Med. 9, eaan3968 (2017).

29. J. Balog et al., Intraoperative Tissue Identification Using Rapid Evaporative Ionization Mass Spectrometry. Sci. Transl. Med. 5, 194ra193 (2013).

30. W. Stummer et al., Fluorescence-guided surgery with 5-aminolevulinic acid for resection of malignant glioma: a randomised controlled multicentre phase III trial. Lancet Oncol 7, 392–401 (2006).

31. K. C. Schafer et al., In Situ, Real-Time Identification of Biological Tissues by Ultraviolet and Infrared Laser Desorption Ionization Mass Spectrometry. Anal. Chem. 83, 1632–1640 (2011).

32. J. Balog et al., In Vivo Endoscopic Tissue Identification by Rapid Evaporative Ionization Mass Spectrometry (REIMS). Angew. Chem. Int. Ed. 54, 11059–11062 (2015).

33. M. F. Keating et al., Integrating the MasSpec Pen to the da Vinci Surgical System for In Vivo Tissue Analysis during a Robotic Assisted Porcine Surgery. Anal Chem 10.1021/acs.analchem.0c02037 (2020).

34. A. M. Porcari et al., Multicenter Study Using Desorption-Electrospray-Ionization-Mass-Spectrometry Imaging for Breast-Cancer Diagnosis. Anal. Chem. 90, 11324–11332 (2018).

35. S. Perwaiz, B. Tuchweber, D. Mignault, T. Gilat, I. M. Yousef, Determination of bile acids in biological fluids by liquid chromatography-electrospray tandem mass spectrometry. J. Lipid Res. 42, 114–119 (2001).

36. C. L. Feider et al., Molecular Imaging of Endometriosis Tissues using Desorption Electrospray Ionization Mass Spectrometry. Sci. Rep. 9, 15690 (2019).

37. M. Behne, H. J. Wilke, S. Harder, Clinical pharmacokinetics of sevoflurane. Clin. Pharmacokinet. 36, 13–26 (1999).

38. J. M. East, C. S. P. Valentine, E. Kanchev, G. O. Blake, Sentinel lymph node biopsy for breast cancer using methylene blue dye manifests a short learning curve among experienced surgeons: a prospective tabular cumulative sum (CUSUM) analysis. BMC Surg. 9, 2 (2009).

39. I. Ali, W. A. Wani, A. Haque, K. Saleem, Glutamic acid and its derivatives: candidates for rational design of anticancer drugs. Future Med. Chem. 5, 961–978 (2013).

40. N. Shenoy, E. Creagan, T. Witzig, M. Levine, Ascorbic Acid in Cancer Treatment: Let the Phoenix Fly. Cancer Cell 34, 700–706 (2018).

41. I. S. Harris et al., Glutathione and thioredoxin antioxidant pathways synergize to drive cancer initiation and progression. Cancer Cell 27, 211–222 (2015).

42. D. Y. Zhang, S. Singhal, J. Y. K. Lee, Optical Principles of Fluorescence-Guided Brain Tumor Surgery: A Practical Primer for the Neurosurgeon. Neurosurgery 85, 312–324 (2019).

43. J. C. O. Richter, N. Haj-Hosseini, M. Hallbeck, K. Wardell, Combination of hand-held probe and microscopy for fluorescence guided surgery in the brain tumor marginal zone. Photodiagn Photodyn 18, 185–192 (2017).

## Supplementary References

1. J. L. Zhang et al., Nondestructive tissue analysis for ex vivo and in vivo cancer diagnosis using a handheld mass spectrometry system. Sci. Transl. Med. 9 (2017).

2. L. S. Eberlin, C. R. Ferreira, A. L. Dill, D. R. Ifa, R. G. Cooks, Desorption electrospray ionization mass spectrometry for lipid characterization and biological tissue imaging. Bba-Mol Cell Biol L 1811, 946–960 (2011).

3. C. L. Feider et al., Molecular Imaging of Endometriosis Tissues using Desorption Electrospray Ionization Mass Spectrometry. Sci. Rep. 9, 15690 (2019).

4. M. Behne, H. J. Wilke, S. Harder, Clinical pharmacokinetics of sevoflurane. Clin. Pharmacokinet. 36, 13–26 (1999).

